# IVNS1ABP Deficiency Disrupts Actin Filament Organization and Leads to Cellular Senescence in a Newly Identified Progeroid Neuropathy Syndrome

**DOI:** 10.1101/2024.10.12.24315363

**Authors:** Fang Yuan, Ye Sing Tan, Haofei Wang, Ain Nur Ali, Qiang Yuan, Shu-Min Chou, Yu-Hsin Yen, Gunaseelan Narayanan, Lei Zhou, Mohammad Shboul, Carine Bonnard, Bruno Reversade, Su-Chun Zhang

## Abstract

A homozygous variant in *IVNS1ABP* was identified in three siblings, displaying progeroid features with severe neuropathy. By generating isogenic induced pluripotent stem cells (iPSCs) from the patients’ fibroblasts and differentiating the iPSCs into neural progenitor cells (NPCs), we found that mutant IVNS1ABP fibroblasts, iPSCs, and NPCs exhibited disrupted cytokinesis, DNA damage and cellular senescence. Correspondingly, cerebral organoids displayed premature differentiation of NPCs to neurons. Molecular profiling as well as biochemical and cellular analysis revealed altered binding of mutant IVNS1ABP to actin /actin-associated proteins and dysregulated actin dynamics during cytokinesis. Taken together, we propose that mutant IVNS1ABP dysregulates actin polymerization and organization which is at least partly responsible for the cellular senescence phenotypes in this progeroid neuropathy syndrome.

## Introduction

Cellular senescence occurs throughout life, contributing to development or aging.^1^ Mitotic cells undergoing senescence display telomere attrition, DNA damage, increased level of senescence-associated β-galactosidase (SA-β-gal), dysregulated metabolism, accumulation of macromolecule aggregates, and senescence-associated secretory phenotypes (SASP).^2^ The causes of cellular senescence are diverse, resulting in highly heterogeneous and dynamic senescence outcomes.

During development, mitotic misregulation can lead to the accumulation of genomic abnormalities, aneuploidy, and cytokinesis failure, ultimately resulting in cellular senescence and premature aging.^3–5^ For instance, reduction of the spindle assembly checkpoint (SAC) protein BubR1 causes increased aneuploidy and senescence in mammals.^6^ BubR1 mutant mice showed progeroid features, including short lifespan, cachectic dwarfism, lordokyphosis, cataracts, loss of subcutaneous fat and impaired wound healing.^7^ In Hutchinson-Gilford Progeria Syndrome (HGPS), mutated lamin A produces a truncated protein progerin, which accelerates cellular senescence by forming abnormal nuclear structures, disrupting cell division and inducing telomere dysfunction.^8,9^ The expression of progerin is also detected during normal aging process,^10^ suggesting that similarities likely exist between premature aging and natural aging, and that natural aging could be understood by studying premature aging cases. However, segmental progeria syndromes do not fully capture the aging process as cognitive functions are often well-preserved in these conditions.^11,12^

We identified a new progeroid syndrome in a family whose teenaged members displayed symptoms like dyschromatosis, whitening hairs, and progressive motor deficits as well as neurological and intellectual deficits.^13^ Exome sequencing coupled to homozygosity mapping revealed a germline homozygous variant in the *IVNS1ABP* gene, which encodes IVNS1ABP, an influenza virus non-structural protein-1 binding protein. It is mainly known for mediating RNA splicing and mRNA export during influenza virus transfection process.^14,15^ Structurally, IVNS1ABP is a Kelch protein, belonging to the KLHL (Kelch-like) subfamily of proteins that function as adaptors for the E3 ligases. Unlike other KLHL proteins, which bind Culin3, IVNS1ABP appears to regulate the proteasome system indirectly through KLHL20.^16^ Moreover, the protein has been found to stabilize F-actin organization in a fibroblast cell line by binding to actin through its Kelch repeats.^17,18^ It is also postulated as a risk gene for primary immunodeficiency in a recent WGS study.^19^ However, IVNS1ABP has not been associated with aging so far, and its role in premature aging and neuropathy is unknown.

Here, we acquired dermal fibroblasts from the patients and their family members, generated isogenic iPSCs from the fibroblasts and differentiated the isogenic iPSCs to neural progenitor cells (NPCs). We found that the fibroblasts, iPSCs, and NPCs with the IVNS1ABP mutation display signature cellular senescent phenotypes. They grew more slowly with an extended cell cycle as well as disrupted cytokinesis and mitotic failure. Proteomics revealed reduced binding of mutant IVNS1ABP to actin and actin-binding proteins, leading to impaired actin polymerization. We propose that dysregulated actin polymerization and organization result in cellular senescence and premature neural differentiation in the progeroid neuropathy.

## Results

### Mutant IVNS1ABP NPCs display increased DNA damage and cellular senescence

Since no model systems were available for the newly identified disease, we generated three iPSC cell lines from dermal fibroblasts of the siblings, two from siblings 1 and 4 carrying a homozygous missense p.(Phe253Cys) variant at an IVR (intervening region) between the BACK domain and Kelch domain of IVNS1ABP and one from the non-affected sibling 2 **(Fig. 1A)**. To investigate the potential roles of IVNS1ABP and reduce the variations derived from different genetic backgrounds, we established three isogenic iPSC pairs by CRISPR/Cas9 **(Fig. 1B & Fig.S1A)**. Pair I (Ctrl_I & MT_I) and Pair III (Ctrl_III & MT_III) were generated from two patient siblings by correcting the mutation site and Pair II (Ctrl_II & MT_II) were generated from the healthy sibling by knocking in the same mutation **(Fig. 1B & Fig.S1A)**. Additionally, we generated an IVNS1ABP knockout (KO) line in Pair III **(Fig. 1B, S1B & S1C)**. The identity and authenticity of these iPSC lines were verified by PCR, Sanger sequencing, off-target analysis and karyotyping as well as the expression of pluripotency markers NANOG and TRA-1-60 **(Fig. S1D)**.

**Figure 1.**
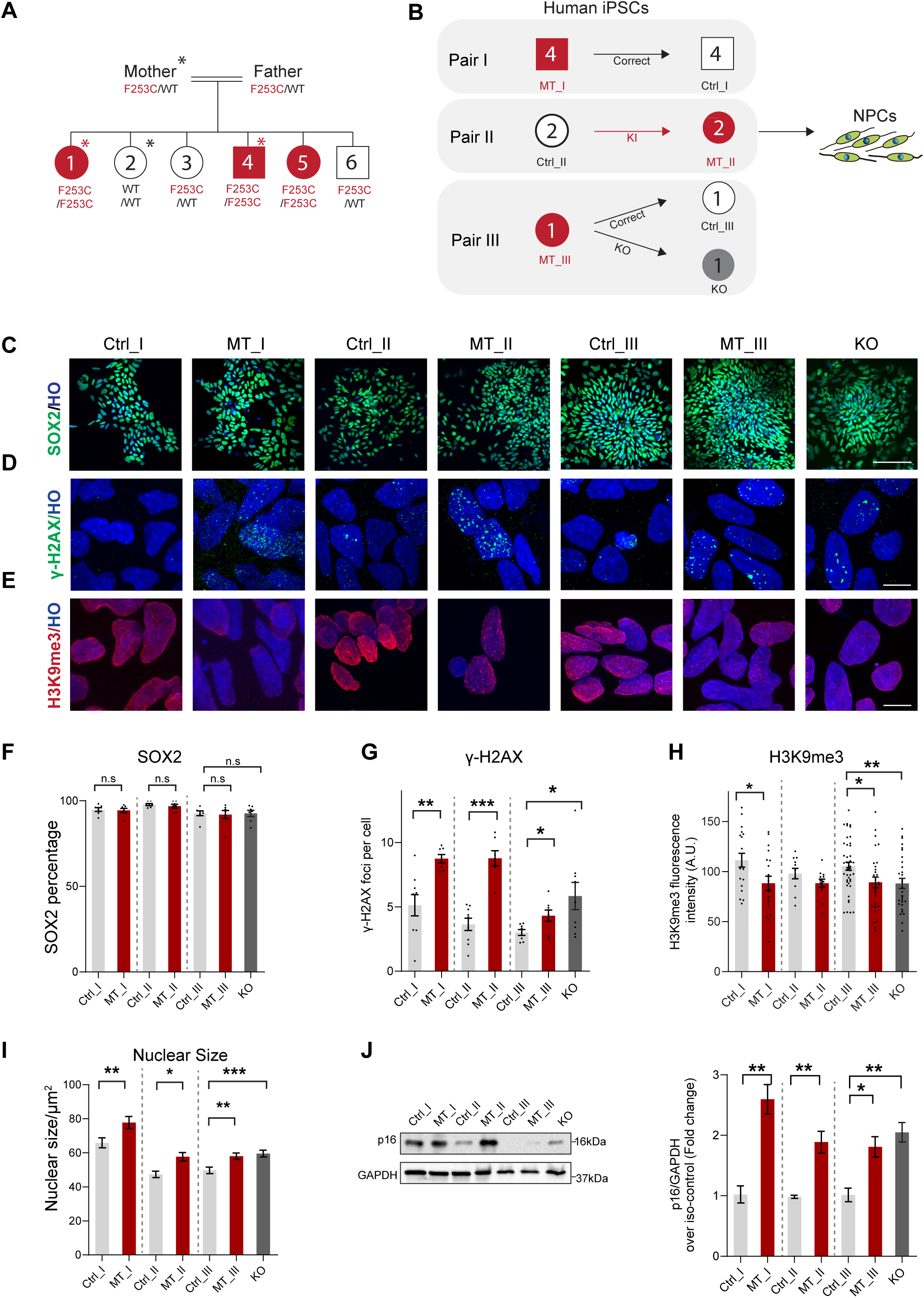
MT IVNS1ABP NPCs display DNA damage and cellular senescence. A. Family tree of the mutation in *IVNS1ABP*. Red color indicates affected patients. * marks the individual origin of primary fibroblasts, including heterozygous healthy mother, one non-affected healthy sibling and two homozygous mutation siblings. B. Graphic flow for disease modeling. Three isogenic iPSCs were generated from one non-affected healthy sibling and two homozygous mutation siblings were reprogramed to iPSCs and then the isogenic lines as well as IVNS1ABP KO isogenic iPSCs were generated. Number on the individual indicates the sibling referred to in Family Tree Graph A. C. Representative images of SOX2+ NPCs derived from all three isogenic pairs of iPSCs. Scale bar, 100μm. D & E. Representative images of γ-H2AX (D) and H3K9me3 (E) staining in NPCs. Scale bar, 10μm. F. Quantification of SOX2+ iPSC-derived NPCs in C. Cells were quantified at day 10 to evaluate the neural lineage differentiation potential. Total cell numbers in each group ∼1000 from 3 independent cultures. n.s.; no significant difference (One-way ANOVA). G & H. Quantification of γ-H2AX foci (D) and H3K9me3 intensity (E). Total cell numbers in each group ∼100 from 3 independent cultures. *, p≤0.05; **, p≤0.01; ***, p≤0.001 (One-way ANOVA). I. Quantification of nuclear size in iPSC-derived NPCs. Total cell numbers in each group ∼80 from 3 independent cultures. *, p≤0.05; **, p≤0.01; ***, p≤0.001 (One-way ANOVA). J. Immunoblot and quantification for p16 in NPCs derived from isogenic pair I, II & III. n≥3. *, p≤0.05; **, p≤0.01 (One-way ANOVA).

Since the patients showed severe neurological and cognitive symptoms as well as premature aging signs, we differentiated the isogenic pairs of iPSCs to forebrain neural progenitor cells (NPCs), following our established protocol.^20^ The identity of NPCs was confirmed by their positive immunostaining for the neural progenitor marker SOX2 **(Fig. 1C)**. Quantitative analysis showed a similar proportion of SOX2+ NPCs between the mutant and isogenic groups at day 10 **(Fig. 1F)**, demonstrating that the neural differentiation capacity at this early developmental stage is not affected by the IVNS1ABP^F253C/F253C^ mutation.

Cellular senescence is often caused or accompanied by DNA damage.^21^ Consistent with this, both IVNS1ABP^F253C/F253C^ mutant (MT) and IVNS1ABP^KO/KO^ (KO) NPCs showed more γ-H2AX foci, as compared to the isogenic controls **(Fig. 1D & 1G)**. H3K9me3, a heterochromatin marker of genome stability, showed decreased expression in MT and KO NPCs as compared to the control groups **(Fig. 1E & H)**. In addition, the nuclear content of the IVNS1ABP MT cells was increased **(Fig. 1I)**. The expression level of CDKN2A/p16, a cell cycle repressor and signature cellular senescence marker, was increased in the IVNS1ABP^F253C/F253C^ NPCs compared to the isogenic controls **(Fig. 1J)**. Taken together, our results demonstrate that IVNS1ABP MT NPCs undergo marked cellular senescence.

### Transcriptomics reveals mitotic misregulation in IVNS1ABP MT NPCs

Senescence phenotypes are highly heterogeneous and dynamic. To systematically explore the cellular senescence caused by IVNS1ABP^F253C/F253C^, RNA-sequencing (RNA-seq) was performed with different cell types (iPSC, NPC) from isogenic pair III (Ctrl, MT and KO) (all with triplicates, 18 samples total) **(Fig. 2A)**. Principal component analysis (PCA) showed that NPC samples were clustered separately from the iPSC samples, suggesting cell type-specific gene expression differences **(Fig. 2B)**. The iPSC samples were clustered together whereas the MT/KO NPCs were segregated from the control NPCs (**Fig. 2B**), suggesting that the impact of MT IVNS1ABP is primarily on somatic cells. Moreover, MT NPC samples clustered close to KO NPC samples, indicating transcriptomic similarities between MT and KO. Similar to the PCA plot, sample to sample correlation analysis showed that MT NPCs presented a close correlation with KO NPCs **(Fig. 2C)**, suggesting that the mutation behaves as a loss-of-function allele.

**Figure 2.**
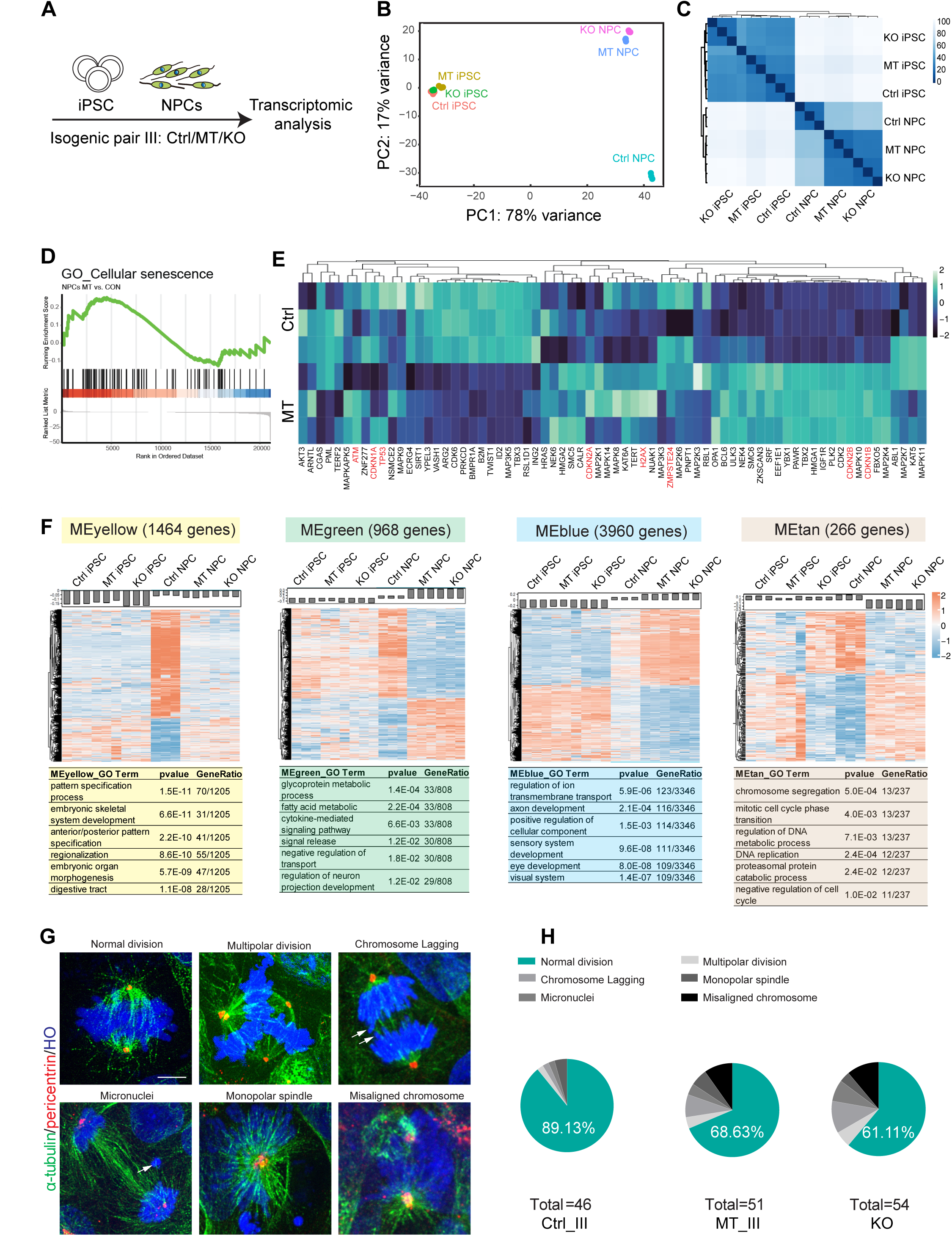
Mitotic misregulation in MT NPCs by RNA-sequencing and immunostaining. A. Schematic flow chart of transcriptomics analysis. B. Principal component analysis (PCA) plot showing sample clustering and variance. C. A heat map showing sample-to-sample distances by hierarchical clustering using DESeq2 based on Poisson distance (blue signifies a high correlation). D. GSEA plots showed Cellular senescence in NPCs, enrichment score = 0.89. E. Heatmap depicting cellular senescence related gene expression in Ctrl and MT NPCs, genes in red are senescence core genes. F. Heat map and GO analysis of eigengenes in the top 4 modules. G. Characterization of mitotic spindle defects in MT/KO NPCs. Metaphase cells were stained with α-tubulin (spindle) and pericentrin (centrosome). White arrows point to the micronuclei between the two daughter cells. Scale bar, 5μm. H. Quantification showing the percentage of abnormal division, including multipolar division; chromosome lagging; micronuclei; monopolar spindle and misaligned chromosome, respectively.

To assess the senescence phenotypes in a comprehensive manner, we performed Gene Enrichment Analysis (GSEA) with Gene Ontology term “Cellular senescence”, which showed higher expression levels of genes in MT NPCs than Ctrl NPCs **(Fig. 2D)**. Among these upregulated genes were some core senescence genes such as *TP53*, *CDKN1A*, *CDK2A*, *CDKN1B*, and *CDKN2B* that are mainly related to cell cycles **(Fig. 2E)**.

We then performed weighted gene co-expression network analysis (WGCNA) to investigate gene sets that are similarly affected by the *IVNS1ABP* mutation. Module-trait analysis of WGCNA identified 39 co-expression modules, of which 9 modules showed differentially-correlated gene expression patterns between WT NPCs and MT NPCs (p<10^-7^) **(Fig. 2F, S2A & S2B)**. The top 4 module eigengenes (MEyellow, MEgreen, MEblue and MEtan) were processed by GO analysis. The yellow module showed enrichment in embryonic development-related terms, while the blue and green modules were enriched in neuron-related terms and metabolism-related terms, respectively **(Fig. 2F)**. Of note, the tan module showed differentiated expression pattern both in iPSC stage and NPC stage. Interestingly, tan module was enriched in mitotic-related terms, including chromosome segregation (GO: 0007059) and mitotic cell cycle phase transition (GO: 0044772) **(Fig. 2F)**, corresponding to the senescence phenotypes **(Fig. 1)** and suggesting cell cycle dysregulation as a potential underlying cause.

At the cellular level, immunostaining for α-Tubulin (mitotic spindle marker) and pericentrin (centrosomes marker), revealed several cell division abnormalities, including multipolar division, micronuclei, lagged chromosomes, monopolar spindle and misaligned spindles **(Fig. 2G)**. The incidence of abnormal divisions in the IVNS1ABP MT and KO cells were significantly higher than the isogenic control group **(Fig. 2H)**, indicating mitotic failure in IVNS1ABP MT NPCs.

### Cell cycle arrest mediating IVNS1ABP mutation induced Mitotic Senescence

The change in transcriptional profiles of cell cycle regulators and mis-alignment of chromosomes during mitosis suggest cell cycle arrest as a mediator of cellular senescence in IVNS1ABP MT cells. Consistent with this, we observed that the patient’s fibroblasts, iPSCs, and NPCs grew more slowly than the healthy cells **(Fig. S3A)**, which is likely caused by reduced proliferation and/or increased cell death. Using cell proliferation assay, with 18h pulse of 5-Ethynyl-2’-deoxyuridine (EdU) to label S phase cells, we showed a reduced proportion of EdU-positive cells in the IVNS1ABP MT fibroblasts as compared to the healthy controls **(Fig. 3A & 3B)**. The expression of Ki67, a marker for proliferation, also decreased in patient fibroblasts **(Fig. 3A, 3B & S3B)**.

**Figure 3.**
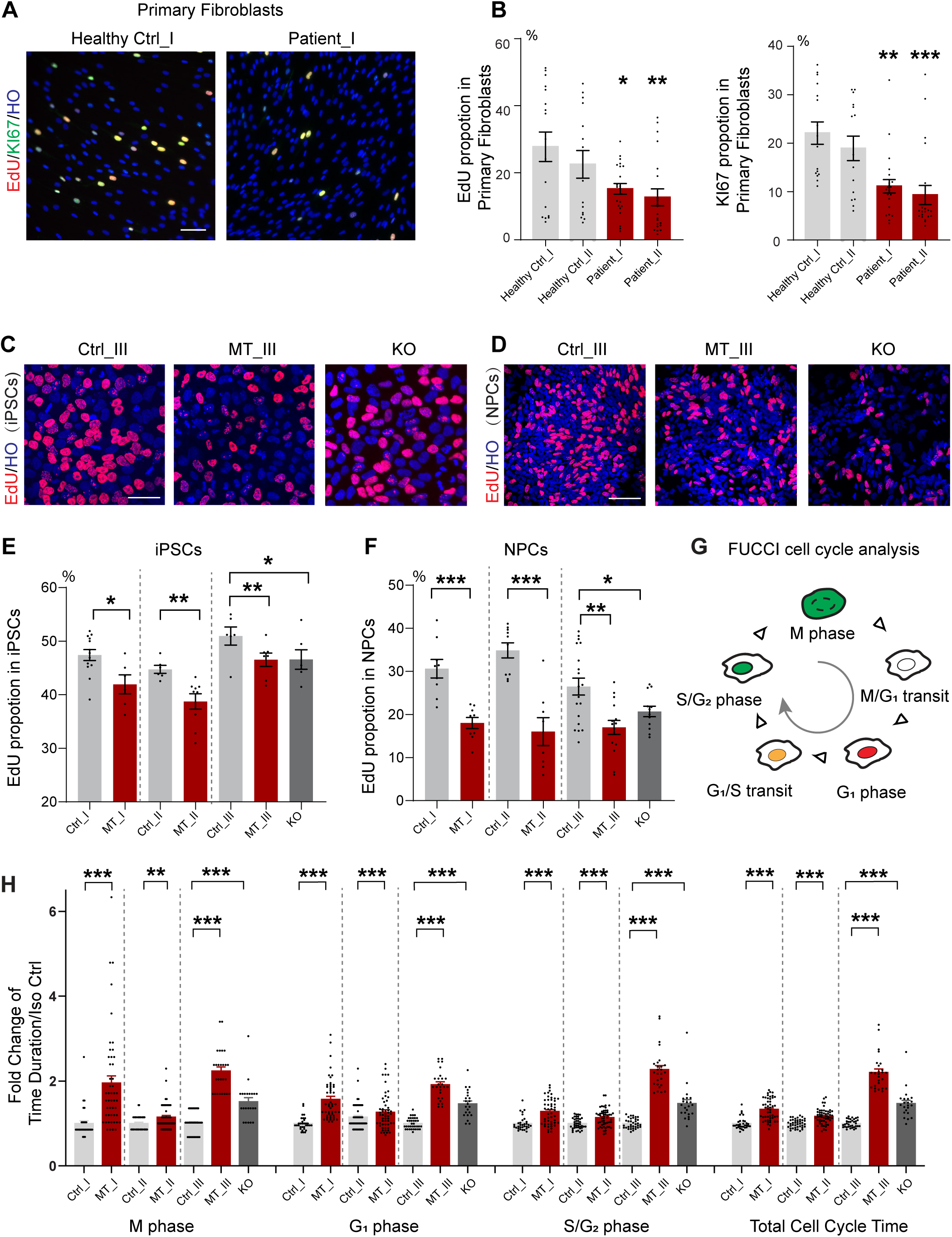
Cell cycle arrest mediating mitotic senescence across multiple IVNS1ABP MT cell lineages. A & B. Representative images of EdU/KI67 staining in primary fibroblasts (A) and their quantification (B). Total cell numbers in each group ∼2000 from 3 independent cultures. *, p≤0.05; **, p≤0.01; ***, p≤0.001 (One-way ANOVA). Scale bar, 100μm. C-F. EdU staining in iPSCs (C) and NPCs (D) from isogenic pair III, quantification in E (iPSCs) and F (NPCs). Total cell numbers in each group ∼1000 from 3 independent cultures. *, p≤0.05; **, p≤0.01; ***, p≤0.001 (One-way ANOVA). Scale bar, 50μm. G. Schematic graph showing time-lapse recording of FUCCI-O labelled iPSCs in different cell cycle stages. H. Quantification of different cell cycle durations with three isogenic pairs, M phase, G_1_ phase, S and G_2_ phase, and total cell cycle duration. Total cell numbers in each group ∼30 from 3 independent cultures; each dot represents one cell. *, p≤0.05; **, p≤0.01; ***, p≤0.001 (One-way ANOVA).

Using isogenic pairs for rigorous comparison, we found that the proportion of EdU-positive cells in the patient and KO iPSCs was significantly lower **(Fig. 3C, 3E & S3C)**, consistent with our observation that the patient iPSCs formed smaller colonies and took longer time to reach confluency. NPCs also showed a decreased proportion of EdU-positive cells in the MT and KO groups compared with their isogenic groups **(Fig. 3D & 3F)**. These results demonstrate that the reduced proliferation is a general defect of IVNS1ABP mutation, with correction of the mutation restoring the cell proliferation capacity. Detection of cell death by cleaved caspase 3 staining showed an increased proportion of positive cells in IVNS1ABP^F253C/F253C^ MT NPCs **(Fig. S4A)**. Thus, the retarded growth is general to many cell types and is caused by both reduced proliferation and increased cell death.

To gain mechanistic insights into cell proliferation reduction, we examined the cell cycle program of iPSCs using the FUCCI system with live cell imaging. FUCCI, a Fluorescent Ubiquitination-based Cell Cycle Indicator,^22^ reports the G_1_ and S/G2 phases by red and green fluorescent reporters in the nucleus, respectively, and the M phase by nuclear envelope breakdown and morphological changes **(Fig. 3G)**. The iPSCs were chosen for FUCCI assay as they are highly homogeneous and possess a relatively shorter division time (16-18hr).^23^ With time lapse recording, we found that the whole cell cycle duration became longer in IVNS1ABP MT cells than in the isogenic control cells (Isogenic pair I: 26h Ctrl vs. 34h MT; Isogenic pair II: 13h Ctrl vs. 16h MT; Isogenic pair III: 13h Ctrl vs. 29h MT) **(Fig. S4)**. Almost all the cell cycle phases were prolonged in the IVNS1ABP MT and KO cells **(Fig. 3H-K)**. Cell death was also observed during live cell imaging of the FUCCI assay, wherein some cells underwent uneven cell division, followed by shrinkage and disappearance of the daughter cells **(Fig. S5C)**. These mitotic failures explain the increased cell death revealed by caspase 3 staining **(Fig. S5A & S5B)**. These results suggest mitotic misregulation as a crucial cause for MT IVNS1ABP-induced cell cycle arrest, DNA damage, and cellular senescence.

### Cell cycle dysregulation is accompanied by premature differentiation in MT IVNS1ABP cerebral organoids

Our patients display severe neurological symptoms and intellectual delay, suggesting the impact of MT IVNS1ABP on neural development and/or maintenance. The cell cycle dysregulation in NPCs would impair developmental programs such as proliferation and/or differentiation. To gain insights into the role of IVNS1ABP in regulating neural development, we generated cerebral organoids from isogenic iPSC lines in addition to 2D cultures of NPCs **(Fig. 4A)**. The measurement of organoid size at 5 weeks showed an apparent reduction in MT organoid size, compared to its isogenic control **(Fig. 4B, 4C)**. In both MT and isogenic control organoids, SOX2-expressing cells around the lumen divided and radially lined up, resembling the ventricular zone (VZ) of the developing cerebral cortex **(Fig. 4B)**. Similar to our observations in 2D cultures of NPCs **(Fig. 4F)**, both SOX2 and Ki67 proportions were reduced in MT organoids compared to isogenic control organoids **(Fig. 4D & 4E)**, indicating that the NPC exits the cell cycle earlier in IVNS1ABP MT organoids.

**Figure 4.**
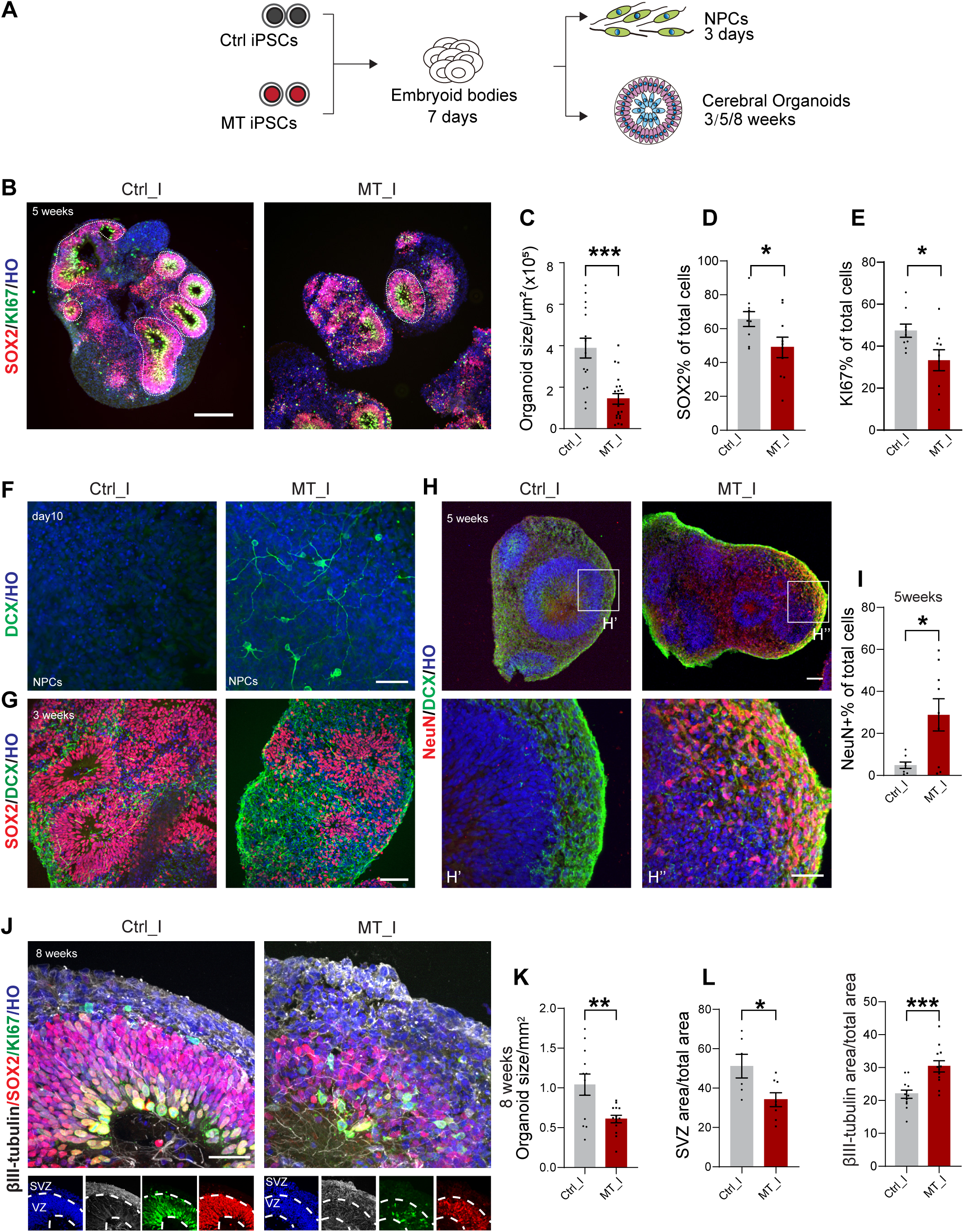
Neural development in 2D and cerebral organoids altered by IVNS1ABP mutation. A. Schematic illustration of monolayer (2D) and cerebral organoid (3D) culture systems. B. Representative images of SOX2 and KI67 staining in 5-week-old cerebral organoids. Scale bar, 100μm. C. Quantification of 5-week-old cerebral organoid size (n = 16 & 20 respectively). D. Quantification of SOX2 percentage in cerebral organoids (n = 10 in each group). E. Quantification of KI67 percentage in cerebral organoids (n=10 in each group). F. DCX staining showing new-born neurons in the MT but not the isogenic control group at day 10 of monolayer cultures. Scale bar, 50μm. G. Representative images of SOX2 and DCX staining to separate neural progenitors from neurons in 3-week-old organoids. Scale bar, 50μm. H. Representative images of organoids stained for DCX and NeuN. The boxed areas are magnified in H’ for the Ctrl organoids and H’’ for the MT organoids. Scale bar, 50μm. I. Quantification of NeuN+ cell percentage in 5-week-old cerebral organoids (n= 8 & 9 respectively). J. Representative images of SOX2, KI67 and βIII-tubulin staining in 8-week-old cerebral organoids, white dash lines indicate VZ and SVZ like structure. Scale bar, 50μm. K. Quantification of 8-week-old cerebral organoid size (n=14 & 12 respectively). L. Quantification of the ratio of SVZ area to total area at 8 weeks (n=6 & 8 respectively). M. Quantification of the ratio of βIII-tubulin covered area to total area at 8 weeks (n=12 in each group). Each dot represents one organoid. *, p≤0.05; **, p≤0.01; ***, p≤0.001 (One-way ANOVA).

Earlier cell cycle exit would alter the cell differentiation program. Indeed, in a 2D neural differentiation culture system, we found early appearance of DCX+ neurons in MT NPCs at day 10, which normally occurs two weeks after differentiation in normal iPSCs **(Fig. 4F)**. This “premature differentiation” indicates a shift in the developmental program. In cerebral organoids, NPCs differentiate to postmitotic neurons following their intrinsic temporal course, presenting a platform to assess if the neural development is altered by MT IVNS1ABP. Indeed, the area covered by DCX+ neurons and neurites was much larger in the MT organoid than in the isogenic organoid at 3 weeks **(Fig. 4G)**. By 5 weeks, the population of maturing neurons, marked by NeuN, was significantly higher in MT than in the isogenic organoids **(Fig. 4H & 4I)**.

To further confirm the premature shift in neural development, we assessed 8-week-old cerebral organoids. The organoid size was still smaller than its isogenic control **(Fig. 4K)**. At this time point, the organoid structure was relatively clear with SOX2-expressing NPCs representing the VZ area and βIII-tubulin expressing neurons and their neurites representing the SVZ area. The VZ-like area was thinner while the SVZ-like area was thicker in the MT organoids as compared to their isogenic counterparts **(Fig. 4J & 4L).** In particular, the βIII-tubulin positive postmitotic neurons were present at the VZ-like area besides the SVZ-like area in the MT organoids whereas they were present mainly in the SVZ-like area in the isogenic organoids at 8 weeks **(Fig. 4J & 4M)**. These results highlight the “premature” neuronal differentiation likely as a result of MT IVNS1ABP-associated cell cycle dysregulation, at least partly explaining the neurological and intellectual deficits.

### Functional loss as an Actin-binding protein due to IVNS1ABP mutation

IVNS1ABP structurally belongs to the adaptor proteins of E3 ligase family. We hypothesized that the MT IVNS1ABP dysregulates cell cycle through altered protein-protein interactions. Hence, we performed a pull-down assay with an IVNS1ABP antibody, followed by mass spectrometry to identify the potential protein interactors of both wild type (Ctrl) and MT IVNS1ABP **(Fig. 5A)** using NPCs from the isogenic pairs **(Fig. S6A)**. We identified 36 proteins in Ctrl NPCs and 30 proteins in MT NPCs, with log_2_Foldchange≥2 compared to IgG control **(Fig. S6B)**. Moreover, 14 proteins were only enriched in Ctrl NPCs with log_2_Foldchange≥2 compared to MT and 9 proteins were enriched in MT NPCs with log_2_Foldchange≥2 compared to Ctrl **(Fig. S6B, Supplementary Table 1)**. Interestingly, IVNS1ABP expression levels were lower in the mutant NPCs than in the control NPCs in all three isogenic pairs, suggesting that the mutation downregulates its protein expression and potential loss-of-function **(Fig. S6C)**. Since cellular senescence and mitotic misregulation could be a consequence of loss-of-function, we focused on the 14 proteins enriched in Ctrl but not MT NPCs. Notably, 10 out of the 14 proteins belonged to the actin/actin-binding protein family **(Fig. 5B)**. The binding between actin and IVNS1ABP was further validated by Co-IP with a pan-actin antibody **(Fig. S7A)**. Apart from ACTB (Actin) itself, most of these proteins were associated with the actomyosin, including myosin-related proteins, MYL6, MYL6B, MYH9, MYH10 and MYL12B, as well as actin-binding proteins, TPM1, CAPZB, ACTR2 and DBN1, which contribute to actin filament organization. For instance, CAPZB regulates actin filament dynamics as a heterodimeric actin capping protein,^24^ ACTR2 mediates actin polymerization as a major component of the ARP2/3 complex,^25^ and DBN1 (Drebrin1) plays a prominent role in dendritic spines as well as the stabilization of actin filaments.^26^ These results demonstrate that IVNS1ABP mutation causes the defects in its binding to actin/ actin binding proteins.

**Figure 5.**
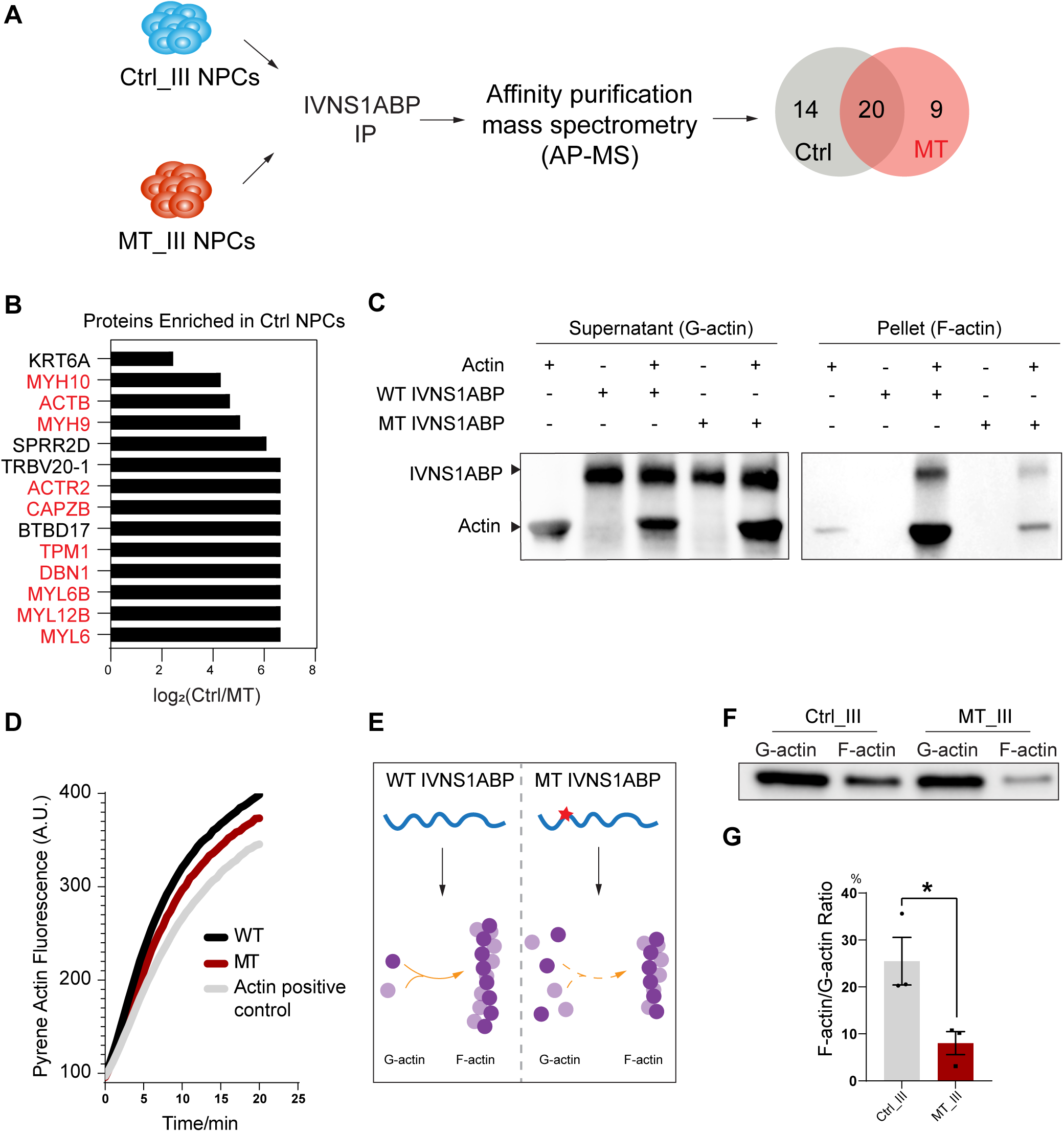
Actin dynamics altered by MT IVNS1ABP. A. Flow chart showing affinity purification mass spectrometry (AP-MS) analysis to identify proteins interacting with WT/MT IVNS1ABP. n=3 in each group (WT, MT, IgG). B. Protein interactors enriched with WT IVNS1ABP, actin/actin binding proteins labelled in red. C. Actin co-sedimentation assay with WT/MT IVNS1ABP as immunoblots showing ACTIN and IVNS1ABP expression. Left blot showing G-actin and IVNS1ABP, right blot showing F-actin spinning down together with IVNS1ABP. D. Pyrene Actin Fluorescence assay showing actin polymerization with WT/MT IVNS1ABP. E. Cartoon showing potential interaction between WT/MT IVNS1ABP and G-actin and F-actin. F. Immunoblot and quantification of F-actin/G-actin ratio in iPSC derived NPCs in isogenic pair III. Data were collected from 3 independent cultures.

The cellular function of actin is regulated by the balance between its monomeric (G-actin) and filamentous (F-actin) forms. To explore the biochemical disparities between WT and MT IVNS1ABP in binding and regulating actin forms, we synthesized WT and MT IVNS1ABP proteins in vitro and conducted an actin co-sedimentation assay. G-actin polymerizes spontaneously, but slowly, under refractory conditions *in vitro* (low salt and low ATP). When incubated with IVNS1ABP, the F-actin pellet was increased in both the WT and MT IVNS1ABP groups, showing the function of IVNS1ABP in facilitating actin polymerization. However, the F-actin amount and binding affinity were significantly reduced in MT IVNS1ABP, compared to WT IVNS1ABP **(Fig. 5C & S7B)**, suggesting a disrupted capability of MT proteins in promoting actin polymerization. This is further confirmed by the pyrene-actin polymerization assay to measure F-actin formation in the presence of the two forms of IVNS1ABP. Indeed, both forms of IVNS1ABP promoted actin polymerization, although the polymerization rate was lower in the presence of MT IVNS1ABP than the WT **(Fig. 5D)**. These findings indicate that the mutation in IVNS1ABP reduces its interaction with F-actin, and thereby dysregulates actin polymerization **(Fig. 5E)**.

Impaired actin polymerization will lead to imbalance between G-actin and F-actin. By using high-speed centrifugation to separate F- and G-actin, followed by Western blotting, we found a reduced ratio of F- over G-actin in MT NPCs compared to the isogenic controls **(Fig. 5F)**. These results suggest that the balance tips towards depolymerization in MT IVNS1ABP cells. We thus evaluated F-/G-actin turnover dynamics in the NPCs using a live actin dye, Sir-Actin. Under the treatment of cytochalasin D (2 μM), which binds to actin filament to prevent actin polymerization, the actin filaments in the MT NPCs began to form aggregates within minutes. Within 30 minutes, the majority of the actin formed aggregates in MT NPCs, whereas the isogenic cells were relatively less sensitive to the concentration of cytochalasin D **(Fig. S8)**, confirming that actin filaments tend to depolymerize in MT IVNS1ABP NPCs. This corresponds to the higher ratio of G-actin over F-actin in unstimulated MT NPCs **(Fig. 5F)**. Taken together, our observations indicate that MT IVNS1ABP fails to balance the polymerization and depolymerization of actin.

### Dysregulated actin turnover contributes to defective cytokinesis and DNA damage in MT IVNS1ABP NPCs

The temporal and spatial regulation of actin dynamics is critical for precise cell division. The altered actin dynamics **(Fig. 5)** and abnormal cell division **(Fig. 2)** observed in MT IVNS1ABP cells suggest a causal correlation between dysregulated actin dynamics and defective cell division. During mitosis, chromosomal segregation is associated with a rounding up of cells and the formation of a contractile actomyosin ring. We found that, F-actin formed a round and even ring to support cell division in Ctrl NPCs synchronized to the G_2_/M phase by nocodazole treatment **(Fig. 6A, B)**. In contrast, F-actin formed an oval and uneven ring in MT NPCs **(Fig. 6A, B)**. Specifically, the actin ring structure was evenly distributed in most of the isogenic cells (89.3%) whereas it was uneven with actin concentrated in some spots, forming dense aggregates in many (24.7%) MT NPCs **(Fig. 6B & 6C)**. Additionally, the MT NPCs displayed a thinner ring cortex **(Fig. 6B & 6D)**. The thin and uneven contractile actin filament ring reduces the rigidity of the cortex during cytokinesis,^27^ contributing to uneven cell division in MT NPCs.

**Figure 6.**
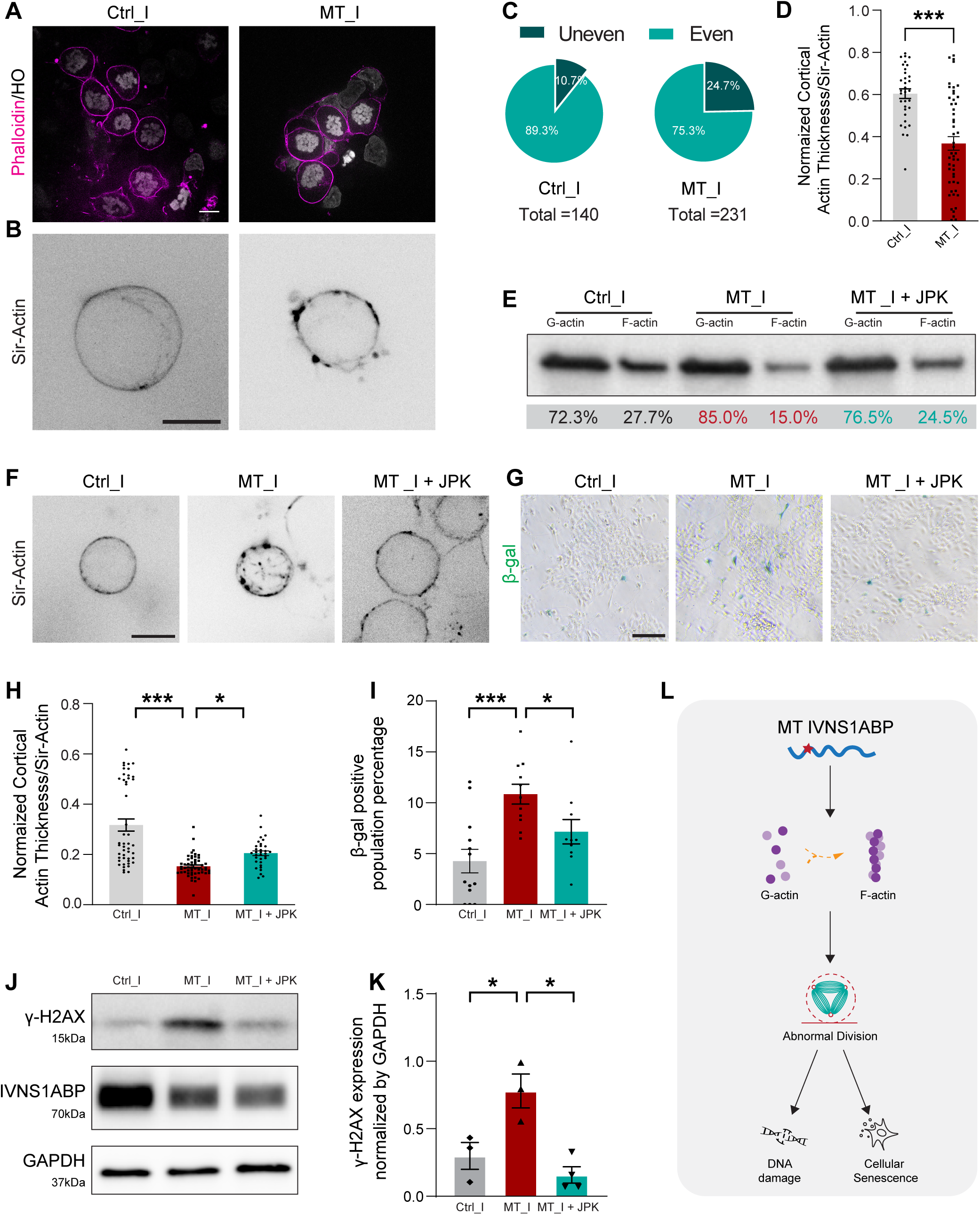
Structure and polymerization of actin altered by MT IVNS1ABP. A. Representative images of actin ring structure during mitosis in NPCs of the MT and isogenic control groups labeled by Phalloidin. Scale bar, 10μm. B. Super-resolution images of live actin ring structure during mitosis in NPCs of the MT and isogenic control groups labeled by Sir-Actin. Scale bar, 10μm. C. Quantification of uneven actin ring structure during mitosis. D. Quantification of normalized actin cortex ring thickness by a self-written MATLab script (n = 33 & 50, respectively). E. Immunoblot of F actin/G-actin ratio in Ctrl, MT and JPK treated MT NPCs. F. Super-resolution images of live actin ring structure during mitosis in NPCs of the MT, the isogenic control groups, and JPK treated MT groups labelled by Sir-Actin. Scale bar, 10μm. G. Quantification of normalized actin cortex ring thickness with JPK treatment (n = 25, 31 and 27, respectively). H. Representative images of β-Galactosidase staining in the Ctrl, MT and JPK treated MT NPCs. Quantification in I (cell numbers ≥ 500 in each group). Scale bar, 50μm. J. Immunoblot and quantification (K) of γ-H2AX, IVNS1ABP and GAPDH in Ctrl, MT and JPK treated MT NPCs. Data were collected from 3 independent cultures. L. Schemic graph showing MT INVS1ABP leads to abnormal division through dysregulated actin dynamics. *, p≤0.05; **, p≤0.01; ***, p≤0.001 (One-way ANOVA).

Given that dysregulated actin dynamics impairs cytokinesis, regulating actin dynamics and rebalancing F-actin/G-actin ratio would, at least partially, restore the actin ring structure and hence cell division. To test this, we applied a low dose of Jasplakinolide (JPK), an actin polymerization drug, to MT NPCs for 24 hours, and found that the F-actin/G-actin ratio was reverted to values similar to those seen in the isogenic control cells **(Fig. 6E)**. Morphologically, the cortical actin ring thickness increased though the aggregates (21.2%, 14/66) did not completely disappear after JPK treatment **(Fig. 6F & 6G)**. Correspondingly, the normal division rate rise from 68.63% (35/51) to 77.08% (37/48), suggesting the cytokinesis defects were mitigated by the enhanced actin polymerization.

We speculated that defective cytokinesis is likely the main cause of DNA damage **(Fig. 1)**. Indeed, we observed that the MT NPCs showed less β–gal and γ-H2AX following treatment with JPK **(Fig. 6H-6K)**. Nevertheless, JPK treatment did not alter the expression level of IVNS1ABP **(Fig. 6J)**. Hence, our findings confirm that the mitotic failure and DNA damage seen in IVNS1ABP^F253C/F253C^ cells are mediated by the dysregulated actin dynamics **(Fig. 6L).**

## Discussion

We have revealed the molecular and cellular mechanisms that contribute to the newly identified progeroid neuropathy associated with a recessive loss-of-function mutation in the *IVNS1ABP* gene.^13^ Multiple patient cell types, from fibroblasts to iPSCs and NPCs, exhibited prolonged cell cycles, increased expression of p16, downregulation of heterochromatins, and increased DNA damage. These cellular senescence phenotypes were eliminated when the mutation was corrected, or mimicked when the mutation was introduced into healthy cells. Interestingly, the phenotypes presented by the MT IVNS1ABP cells resembled those by IVNS1ABP KO cells, suggesting a potential loss of function of IVNS1ABP in the disease pathogenesis. Furthermore, proteomics and transcriptomics studies pointed to alterations in actin and its associated proteins, in addition to dysregulation of proteins involved in cellular senescence. Indeed, MT IVNS1ABP showed decreased binding affinity to actin, leading to dysregulated actin dynamics during cytokinesis, and subsequently, premature neural differentiation and cellular senescence. Strikingly, restoration of the F- and G-actin balance, even in the presence of IVNS1ABP mutation, mitigated cellular senescence. Taken together, our study has identified dysregulated actin dynamics caused by MT IVNS1ABP as a mediator of premature aging in this newly identified progeria-like syndrome.

### The biological function of IVNS1ABP

IVNS1ABP was first identified as a novel human protein termed NS1-BP, in the search for interacting proteins of influenza A virus NS1 protein, through a yeast interaction trap system.^14^ The murine homolog was named as Nd1 (*ncx* downstream gene 1), as it was isolated from an *Ncx*-deficient mice. Nd1 encodes two isoforms— a long (Nd1-L) and a short (Nd1-S) form—with the former being the actin-binding isoform.^18^ In human, it has only one reported form under normal condition, equivalent to Nd1-L. Notably, *Nd1*-deficent mice showed no cardiac or gross anatomical abnormality,^28^ although our human cellular IVNS1ABP KO model showed severe deficits in the CNS, highlighting the unique functions of human IVNS1ABP.

As a Kelch protein, IVNS1ABP, when mutated, may contribute to neuropathy and aging via multiple pathways. The highly conserved BTB/BACK domain suggest its recruitment to E3 ubiquitin ligase complex, indicating that IVNS1ABP plays a role in protein homeostasis. We indeed observed protein aggregations and enlarged lysosomes in multiple IVNS1ABP mutated cell types including NPCs and neurons (Carine Bonnard et al., 2024). Mutations in its paralog KLHL16 result in similar outcomes, where patients present with Giant axonal neuropathy-1 (GAN1) due to impaired proteostasis and accumulation of cytoskeleton proteins in axons.^29^ Unlike many other Kelch proteins, IVNS1ABP does not bind Cullin3, but may regulate ubiquitination through interaction with other KLHL family members such as KLHL20.^16^ Other KLHL proteins, such as KLHL1, Mayven (KLHL2) and IPP (KLHL27), have been shown to bind actin directly.^30–32^ Our proteomics analysis and Co-IP further validated the interactions of IVNS1ABP with actin and actin binding proteins, suggesting that IVNS1ABP may regulate cellular activity via actin.

### Mutant IVNS1ABP dysregulates actin polymerization and mitosis

Actin, a central player in basic cell function, is highly conserved. Surprisingly, IVNS1ABP mutation impairs its ability to bind actin, leading to reduced F-actin/G-actin ratio and disorganization of actin filaments, such as their aggregation, during cell division. Furthermore, alterations in F-actin/G-actin ratio in the mutants suggest a role of IVNS1ABP in actin polymerization/depolymerization, and consistent with this, our polymerization assay revealed a tendency of actin filaments to depolymerize in IVNS1ABP MT cells.

A precise regulation of actin polymerization and depolymerization is crucial for many biological processes such as cytokinesis. At the metaphase when the cell prepares to divide, actin polymerizes and the actin filaments organize into a cortical ring that serves as an anchor for microtubules to pull the chromatins to the opposite directions equally.^33^ However, our studies showed that IVNS1ABP mutations altered the dynamics of actin polymerization, resulting in the formation of a thin and uneven cortical ring. We propose that this could be the basis of several kinds of defects during cytokinesis, including multipolar division, micronuclei, lagged chromosomes, monopolar spindle and misaligned spindles. These mitotic catastrophes result in cell death, evidenced by disappearance of cells shortly following division. Severe mitotic failure and prolonged cell cycles create genome instability and DNA damage, contributing to senescence.

### IVNS1ABP mutation leads to mitotic senescence

Mitotic senescence is a common mechanism underlying progeroid syndromes. Similar to the most common progeroid syndrome HGPS, our patient cells exhibit retarded cell growth, readily characterized by the slow expansion of the dermal fibroblasts, smaller colonies of iPSCs, and reduced incorporation of EDU in NPCs. At the molecular level, MT NPCs exhibit upregulated CDK inhibitor CDKN2A/p16, increased DNA damage, loss of chromatin components and increased expression of cellular senescence genes. However, in contrast to HGPS where CNS senescence is absent, patients with the IVNS1ABP mutation display senescence in the CNS cells too. This is likely due to the fact that the Lamin A mutation in HGPS does not affect Lamin C-expressing CNS cells.^34^ In contrast, the IVNS1ABP mutation in this newly identified disease negatively regulates actin, which is universally expressed in all cell types including neural cells. This could explain the reporting of neurological defects, additional to progeria symptoms, in patients with the IVNS1ABP mutation.

Furthermore, dysregulated actin filament organization during cytokinesis results in cell cycle arrest, which can lead to precocious cell cycle exit and “premature” differentiation of neural progenitors. Indeed, the post-mitotic neurons appeared substantially earlier in the mutant cells in both our 2D cultures and 3D organoids. The early cell cycle exit and premature differentiation results in fewer neurons and axons, reflected in the thinner corpus callosum and cerebellar vermis exhibited by the patients. Actin dynamics is crucial for synaptic plasticity in neurons; actin dysregulation is highly associated with age-related cognitive decline,^35,36^ potentially explaining the cognitive impairment in our patients. Dysregulated actin dynamics impairs axonal integrity, additional to impaired proteostasis,^13^ leading to neuropathy. Further studies are needed to examine in detail the impact of MT IVNS1ABP on neural development, maintenance, and plasticity.

In summary, we identified a new premature aging syndrome with neurological and cognitive deficits caused by a homozygous variant in the *IVNS1ABP* gene. MT IVNS1ABP, similar to the gene KO, impairs its ability to bind actin and actin-associated proteins, and thus, dysregulates actin polymerization. Dysregulated actin polymerization disrupts precise cytokinesis in mitotic cells, leading to cell cycle arrest and cellular senescence. Precocious cell cycle exit results in premature neural differentiation, leading to fewer neurons and axons as well as impaired synaptic function, and hence complex syndromes, such as neurological and cognitive deficits. While this study focuses on actin dysregulation by mutant IVNS1ABP, it is likely that other pathways, such as proteostasis,^13^ are involved. Further studies are necessary to define the new disease and to find targeting therapeutics.

## Methods

### Culturing of Human iPSCs and NPCs

Human iPSCs (Passage 10–40) were maintained on vitronectin-coated plates (Life Technologies) with Essential 8 medium, which was changed daily. Cells were passaged every 5 days through ethylenediaminetetraacetic acid (EDTA) (Lonza) digestion. Neuron differentiation was carried out according to our previously established protocol.^20^ Briefly, hPSCs were detached by dispase (Life Technologies) to form embryoid bodies (EBs) and then cultured in neural induction medium (DEMEMF12 medium supplemented with 100x N2 and 100x NEAA). After floating culture for 7 days, EBs were attached, and rosette structures could be observed at day 10–16. At day 16, rosette colonies were detached manually with a 1 ml pipette. Non-neural epithelial clones were removed at this stage. Neural progenitors were used for analysis at day 10 or day 20. To culture organoids, EBs were continuously cultured in neural induction medium (NIM) for 3 weeks or longer.

### Gene Editing of iPSCs by CRISPR/Cas9

Human iPSCs were maintained under feeder-free conditions, and treated with Rho Kinase (ROCK) inhibitor 24 hours before electroporation. A guide RNA was designed using the CRISPR design tool (http://chopchop.cbu.uib.no/); the ∼75nt donor single-stranded oligodeoxynucleotides (ssODNs) were designed with homologous genomic flanking sequence centered around the CRISPR/Cas9 cleavage site containing point mutation.^37^ The iPSC cultures (1 × 10^6^) were dissociated into single cells by EDTA for 5 minutes, and then were mixed with reagents (4 μg Cas9 nuclease, 150 pmole sgRNA and 2 nmole ssODN) (IDT technologies). The cell mixture was electroporated using the Neon transfection System (Invitrogen) with the following parameters: Voltage 1200V, Width 30 ms and 1 Pulse. After electroporation, the cells were reseeded on a vitronectin-coated six-well plate in Essential 8 medium, with the addition of ROCK inhibitor for the first 24 hours. Stable colonies were selected 2-3 days after seeding. Positive colonies were confirmed with Sanger Sequencing, following which they were expanded and stored. The top 5 predicted off-target sites were amplified and Sanger Sequenced, and then compared with the unedited cell line. Karyotype tests were performed by the Cytogenetics Lab at Singapore General Hospital. Pluripotency of the iPSCs were confirmed by TRA-1-60 and NANOG staining. The guide RNA sequence, ssODN template sequence and off-target information are included in **Supplementary Table 2**.

### Immunochemistry

Cells cultured on coverslips or organoid samples were fixed in cold fresh 4% paraformaldehyde for 30 min and rinsed three times with phosphate buffered saline. Organoid samples were further dehydrated in PBS solution containing 30% sucrose for 1 day and sectioned at a 30 μm thickness. Cells or organoid sections were treated with 0.2% TritonX-100 for 10 min and blocked in 10% donkey serum for 1 hr. Cells were incubated at 4°C overnight in primary antibody diluted with 0.1% triton and 5% donkey serum. On the second day, cells were incubated in secondary antibody diluted in 5% donkey serum for 30 min at room temperature. Coverslips were mounted for fluorescent imaging. The primary and secondary antibodies are listed in **Supplementary Table 3**.

### Western Blot

Cells were washed with cold PBS, scratched and lysed on ice using RIPA lysis buffer (Thermo Fisher) together with protease inhibitor, phosphatase inhibitor, phenylmethylsulfonyl fluoride and dithiothreitol. Total protein concentration was measured by BCA protein assay. 4X Laemmli sample buffer was added into the protein lysate and boiled at 100℃ for 5 mins. Protein samples were loaded on 4%-20% Mini-Protein SFX precast gel (Bio-rad), and then transferred to polyvinylidene difluoride membranes, blocked with 5% non-fat dry milk TBST, and then incubated with primary antibodies overnight at 4°C. Signals were visualized using horseradish peroxidase-conjugated secondary antibodies, ECL system and captured with ChemiDoc system. Antibodies are included in **Supplementary Table 3**.

### FUCCI cell cycle analysis

The FUCCI system was introduced into cells through lentiviral transfection. For viral particle generation, HEK293T cells were transfected with second generation lentivirus plasmids pCMV-VSV-G (Addgene #8454), psPAX2 (Addgene #12260) and pBOB-EF1-FastFUCCI-Puro (Addgene #86849). Viral particles were collected 48 hours post transfection and purified by ultracentrifugation. IPSCs were transfected with FUCCI virus and screened by puromycin for two days. FUCCI-expressing cells were then expanded and passaged for subsequent live cell imaging experiments. iPSCs positively expressing FUCCI were seeded on confocal dishes and recorded 48h after passaging.

### Measurement of G-Actin/F-Actin ratio

G-Actin/F-Actin ratio was determined by immunoblotting of a specific actin antibody using a G-actin/F-actin assay kit (Cytoskeleton). Briefly, Ctrl/MT NPCs were lysed in a detergent-based lysis buffer, which stabilizes and maintains the G-and F-forms of cellular actin. The two forms of actin differ in that F-actin is insoluble, whereas G-actin is soluble in the lysis buffer. The G-actin and F-actin fractions were separated using ultracentrifuge (TLA100 rotor; Beckman Coulter) at 100,000g for 1h at 37°C. G-actin was persevered in the supernatant while F-actin in the pellet was depolymerized to G-actin in the same volume of depolymerization buffer. Equal amounts of supernatant (G-actin) and pellet (F-actin) fractions were mixed with 5X SDS sample buffer and processed for western blotting.

### RNA-seq analysis

Human iPSCs (Ctrl/MT/KO) and NPCs (Ctrl/MT/KO) from isogenic pair III were collected for total RNA extraction and each sample was processed in triplicates. Whole transcriptome RNA-seq was performed by Novogene Singapore Pte.Ltd. with 40 million 150bp pair-end reads per library. The quality of the sequencing library was evaluated with FastQC. Following this, Salmon was used to align sequencing reads to the human transcriptome (hg38) ^38^, and Deseq2 was used for differential gene expression analysis. Differentially-expressed genes between MT and WT samples were identified based on p-value ≤ 0.05 and fold-change ≥ 2.^39^ Weighted gene coexpression network analysis (WGCNA) was performed with WGCNA package in R.^40^ Co-expression gene modules between MT and WT were constructed using Blockwise, with default settings and a power threshold of 9. Each resulting module was subsequently subjected to functional analysis using the Modules function. The genomic background for this analysis encompassed all genes expressed in the current dataset. Each module was enriched for GO biological processes using ClusterProfiler.^41^ Only gene sets that passed a multiple test adjustment using the Benjamini– Hochberg procedure (adj. p≤0.05) were deemed significantly-enriched in a biological process.

### Co-Immunoprecipitation

Approximately 20 million Ctrl/MT NPCs were lysed for 30 mins through vortexing and pipetting up and down, using 500μL Pierce™ IP buffer (ThermoFisher) supplemented with protease inhibitors. 25μl Protein G beads (Pierce™ Protein A/G Magnetic Beads, ThermoFisher) were pre-washed three times with Tris-buffered saline containing 0.05% Tween-20, and subsequently collected with a magnetic stand. The beads were further incubated with either IVNS1ABP antibody (Anti-Ms, Santa Cruz) or IgG (Anti-Ms, Millipore) antibody for 1h with gentle rotation at room temperature, followed by an overnight incubation with cell lysates at 4°C. On the next day, the supernatant was saved for analysis and beads were washed thrice with Tris-buffered saline containing 0.05% Tween-20, and once with water. After washing, the supernatant containing the antigen was separated from the beads magnetically by elution. Eluted bead-bound proteins were further processed with neutralizing buffer (1M Tris) before electrophoresis in SDS-PAGE. The total protein lysate (input) and the IP fractions (output) were analysed by immunoblotting using the IVNS1ABP antibody (Anti-Rb, Novus).

### Affinity purification mass spectrometry (AP-MS) analysis

The CO-IP fractions were resolved with lysis buffer and universal nuclease (EasyPep™ Mini MS Sample Prep Kit, ThermoFisher) to extract the proteins. Protein quantitation was done using DC Protein Assay (Bio-Rad Laboratories Inc., California, USA). The protein solutions were then reduced, alkylated, digested and cleaned-up according to manufacturer’s instructions (EasyPep™ Mini MS Sample Prep Kit, ThermoFisher). Peptides were reconstituted in 2% Acetonitrile and 0.1% formic acid in water, and peptide concentration was determined by Thermo Scientific™ Pierce™ Quantitative Fluorescent Peptide Assay.

The reconstituted samples were then analyzed on an EASY-nLC 1200 system coupled to Orbitrap Exploris^TM^ 480 mass spectrometer (ThermoFisher).The Easy-nLC system was equipped with a 100 C18, 3 μm, 75 μm x 2cm in-line trap column of PepMap and a C18, 2 μm, 25cm x 75 μm Easy-spray Pepmap RSLC column. The EASY-nLC was operated at a flowrate of 300nL/min. Mobile phase A consisted of 0.1% formic acid in LC-MS grade water and mobile phase B was made up of 0.1% formic acid and 80% acetonitrile in LC-MS grade water. The gradient comprised of a 29min step gradient from 5% to 60% mobile phase B and 1min from 60% to 98% solvent B. Orbitrap ExplorisTM 480 mass spectrometer was operated in data-independent and positive ionization mode. LC-MS/MS analysis was performed as described below: MS1 spectra were recorded at a resolution of 60k with MaxIT mode set to auto. The scan range was 350 to 1600 m/z for full scan. The automatic gain control (AGC) target was set to custom with normalized AGC target at 300%. Peptides were then selected for ddMS2 using HCD Collision energy at 28% with a fixed collision energy mode and the fragments were detected in the Orbitrap at a resolution of 15k with auto MaxIT. The isolation window was set to custom with m/z set to 1.6. The first mass was set at m/z 120. The AGC target was set to custom with normalized AGC target set to 75%.The resulting MS/MS data were processed using Proteome Discoverer version 2.5 (ThermoFisher).

### Protein Generation and Actin Co-sedimentation Assay

WT/MT IVNS1ABP cDNA fragments were obtained by PCR amplification of the cell extracts, and subsequently inserted into pTNT™ Vector (Promega) following manufacturer’s instructions. The WT/MT IVNS1ABP constructs were then processed using the TNT® Quick Coupled Transcription/Translation Systems (Promega) to express WT/MT IVNS1ABP proteins. The expression of these proteins was further verified by immunoblotting with an IVNS1ABP-specific antibody.

Actin co-sedimentation assay was performed using the actin-binding spin-down kit (Cytoskeleon). Briefly, G-actin protein solution was incubated with WT/MT IVNS1ABP protein at RT for 30mins and sedimented at 150,000 x g (TLA100 rotor; Beckman Coulter) for 1.5h at RT. The G-actin and F-actin fractions were then separated and immunoblotted with actin and IVNS1ABP antibody.

### Pyrene-actin Polymerization Assay

Actin polymerization rates were measured using commercial Actin Polymerization Biochem Kit (Cytoskeleton). The fluorescence signal of pyrene-monomeric actin is enhanced 7-10 times during its assembly into filaments, making it an ideal tool for tracking actin polymerization.^42^ Briefly, WT/MT proteins were adjusted to a concentration of 100nM in TBS buffer. The pyrene G-actin was prepared 1 hour after depolymerization of actin oligomers and included a 30-min ultracentrifuge to avoid F-actin. Blank buffer (5 mM Tris-HCl pH 8.0, 0.2 mM CaCl_2_) was added into a 96-well black plate as a background control. Pyrene G-actin/Pyrene G-actin and target protein mixture with blank buffer were set as a positive control and experimental group, respectively. The plate was pre-incubated in a fluorescence spectrophotometer (Tecan Infinite M200 Microplate Reader) for 3mins to set up the baseline and the reaction was initiated by adding actin polymerization buffer (500 mM KCl, 20 mM MgCl_2_, 10 mM ATP). Fluorescent signals were measured every 30s for 1h with excitation at 365 nm and emission at 385 nm.

### Microscopy/ live cell Imaging

Images were captured using a Nikon Ti2 inverted microscope equipped with Yokogawa spinning disk confocal, GATACA super-resolution systems (SIM) and sCMOS camera (Prime 95B). For live imaging experiments, images were acquired at 37°C with 5% CO^2^ using an on-stage incubator and CO_2_ mixer (LCI), and cells were imaged in multi channels by sequential laser excitations at 488 nm, 561 nm, or 642 nm through a quad-bandpass dichroic mirror (Semrock) and single band emitters (Semrock). For FUCCI recording, movies were acquired on a single z plane with a speed of 5min/frame or 10min/frame, and exposure times in the 100–300ms range. Actin ring structure was captured with SIM microscopy. Immunostaining images were taken with z-stack acquisition with 0.5 or 1⎧m step-size and processed with maximum projection images of multiple z-stacks.

### Quantification and Statistical Analysis

Data was analysed by GraphPad Prism version 8. All bar graphs are presented as mean values ± SEM. Replicate sizes and error bars are indicated in the figure legends. Two-tailed Student’s t tests were performed for datasets with two groups. ANOVA analyses were used for comparisons of data with more than two groups. *, p≤0.05; **, p≤0.01; ***, p≤0.001.

## Acknowledgements

We would like thank all members from Su-Chun’s laboratories for discussions and suggestions. We thank Andrew Petersen for technical help on CRISPR editing. F.Y. was supported by Duke-NUS Medical School Khoo Postdoctoral Fellowship-(KPFA/2020/0038) and a National Medical Research Council Open Fund (OFYIRG22jul-0021). C.B. was supported by a NMRC Open Fund - Young Individual Research Grant (OF-YIRG/0048/2017). B.R is a fellow of the National Research Foundation (NRF, Singapore) and Branco Weiss Foundation (Switzerland) and an EMBO Young Investigator. This work was also funded by a Strategic Positioning Fund for Genetic Orphan Diseases (SPF2012/005) and an inaugural A*STAR Investigatorship from the Agency for Science, Technology and Research in Singapore to B.R; S-C.Z. was supported by Singapore Ministry of Education Research Fund, MOE2018-T2-2-103; Singapore Ministry of Health Research Fund, MOH-000207 and 000212.

## Author contributions

F.Y. conceived and designed the study, performed isogenic cell lines generation, cell differentiation, immunostaining and imaging, immunoblots, data analysis and interpretation, and wrote the manuscript. Y.S.T. performed cell culture, immunostaining and imaging, immunoblots and data analysis. H.W. performed RNA-seq analysis. G.N. generated one pair of isogenic cell line. L.Z. performed AP-MS experiments and proteomic data analysis. A.N.A., M.S., C.B. and B.R. provided the disease diagnosis, patient’s fibroblasts and iPSCs. Q.Y., S-M.Z. and Y-H Y performed data interpretation. S.-C.Z. conceived, designed and supervised the study, and wrote the manuscript. All authors reviewed the manuscript.

## Data and code availability

RNA-sequencing data were deposited to Gene Expression Omnibus (GEO) under GSE270946 and raw proteomics data were deposited to ProteomeXchange (Identifier: PXD053645). These are publically available as of the date of publication. This paper does not report original code.

