## Supplementary Figures for "IVNS1ABP Deficiency Disrupts Actin Filament Organization and Leads to Cellular Senescence in a Newly Identified Progeroid Neuropathy Syndrome"

### Figure S1 Generation of isogenic iPSCs

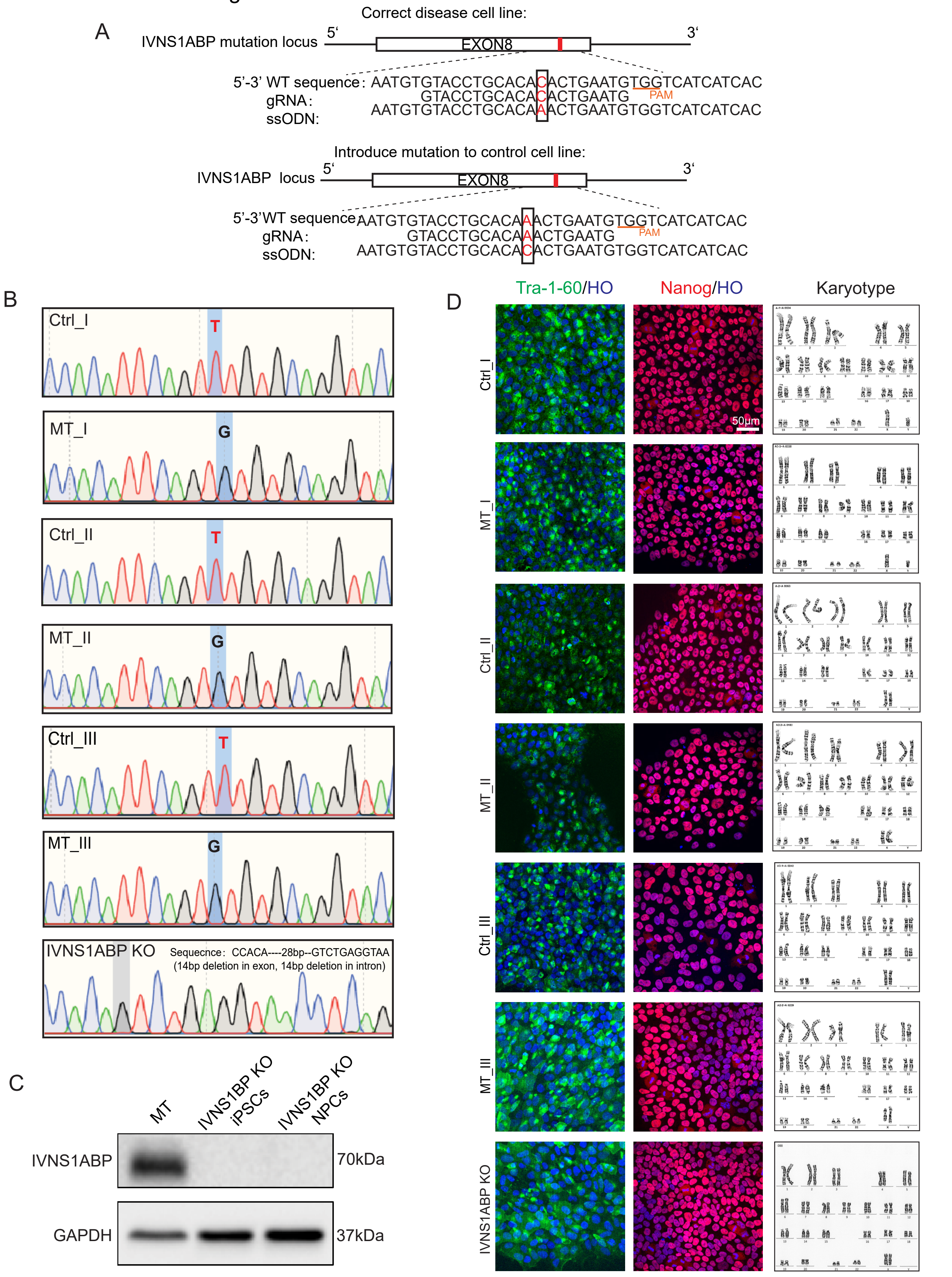


A. Schematic strategy for Crispr editing isogenic iPSC pairs. Upper pannel is to correct the patient IPSCs, while below pannel is to introduce the point mutation into the healthy iPSCs.
B. Sanger sequencing of the DNA fragraments including the mutation site.
C. Immunblot shows the successful knockout of IVNS1ABP
D. Representative images of Tra-1-60 and Nanog and karyotype of the isogenic pairs of iPSC cell lines.

Figure S2 Weighted gene co-expression network analysis of RNA sequencing

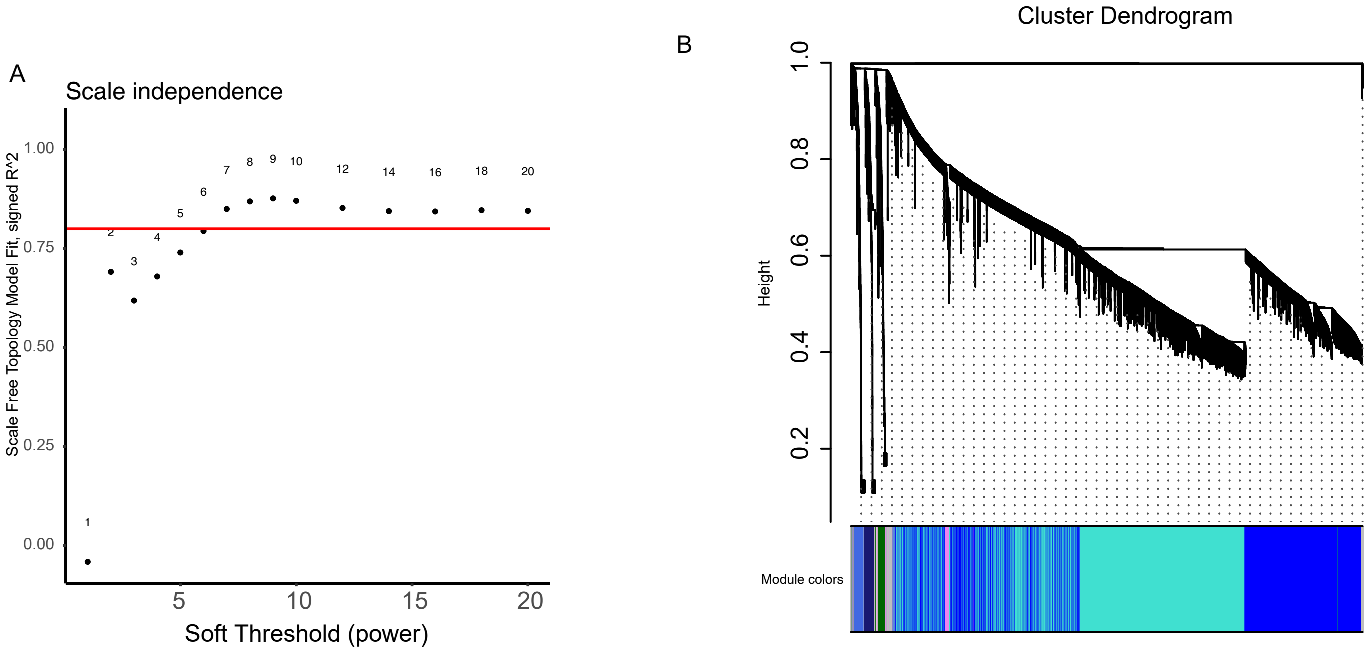


A. The β-power required to satisfy scale-free topology (SFT).
B. Hierarchical gene cluster tree and module structure and gene-module color bands. The color band underneath the tree indicates the detected modules.

Figure S3 Reduced cell prolifreation with IVNS1ABP mutation

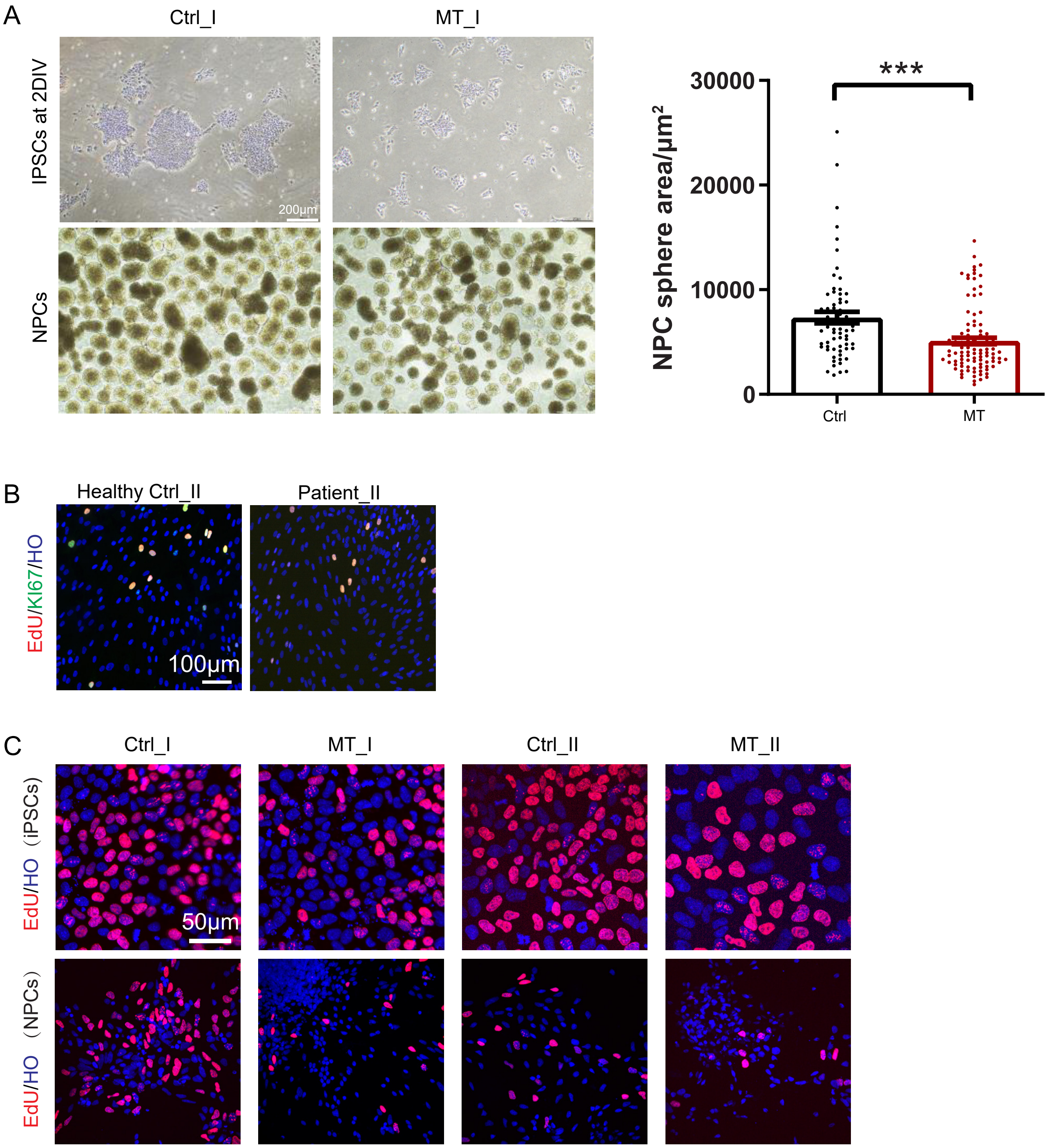


A. Representative bright field images of iPSCs and NPCs from isogenic iPSC pair I. Quantification on the right (n~50 in each group, respectively).
B. Representative images of EdU/KI67 staining in primary fibroblasts. Quantification in Fig. 3B
C. EdU staining in iPSCs and NPCs from isogenic pair I & II. Quantification in Fig. 3E & 3F.

Figure S4 Mitotic misregulation recorded by live cell imaging with FUCCI labeling

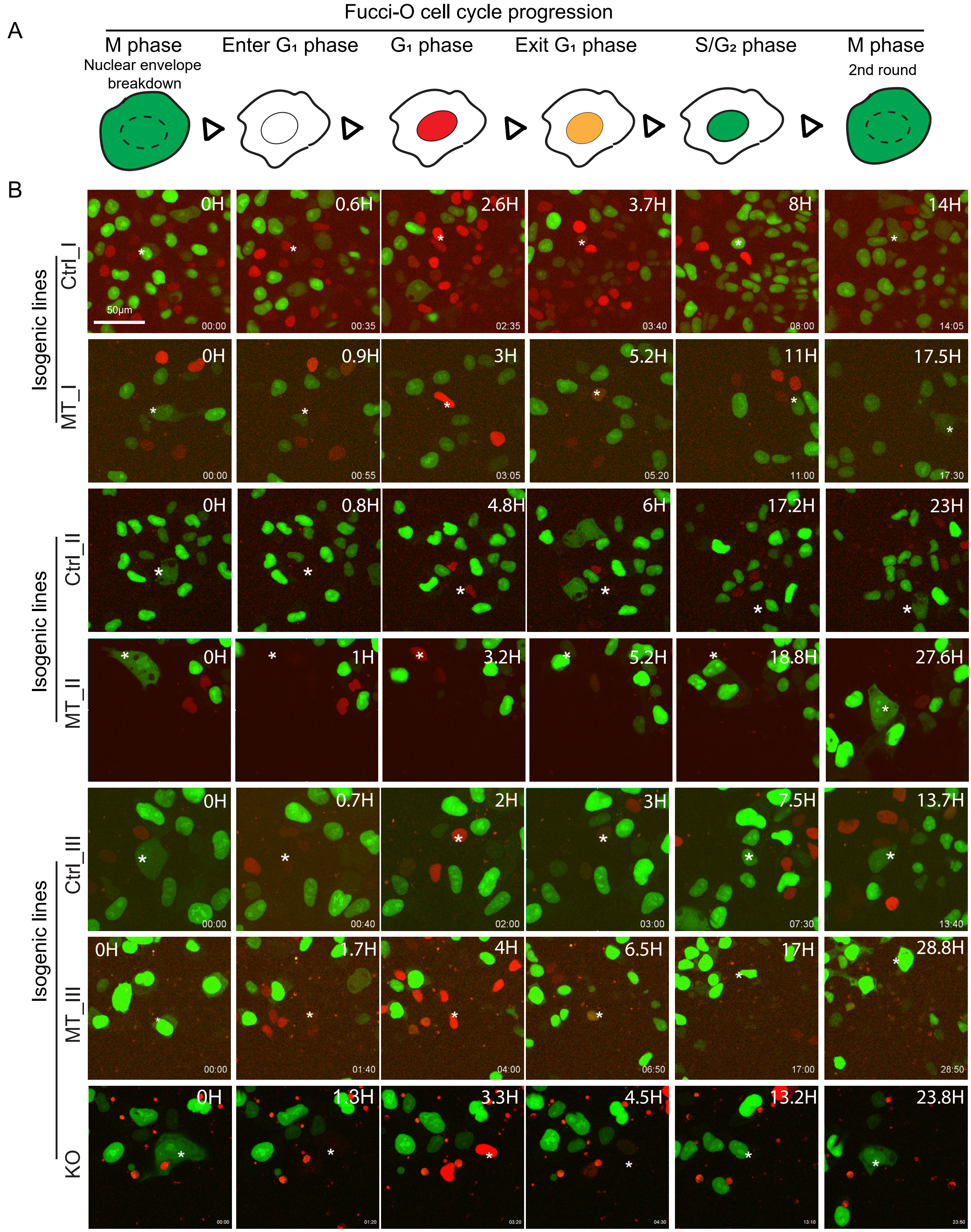


A. A schematic diagram illustrating the color changes of the nucleus in each cell cycle phase, corresponding to the live cell images below.

B. Representative images showing a complete cell division cycle in each group. Cell was labeled by the star mark. The sequence begins with the nuclear envelope breakdown, designated as time 0H, marking the entry into the M phase. The transition from GFP to RFP, indicated by a transparent color, suggests the entry into the $G_1$ phase. RFP in the nucleus confirms the cell is in the $G_1$ phase. The transition from RFP to GFP indicates the cell is entering the $S/G_2$ phase, with GFP expression in the nucleus during this phase. And then the second time of nuclear envelope break down suggested the cell is entering the second round of cell division. Each fragamented time was seperated and recorded.

Figure S5 increased cell death with IVNS1ABP mutation

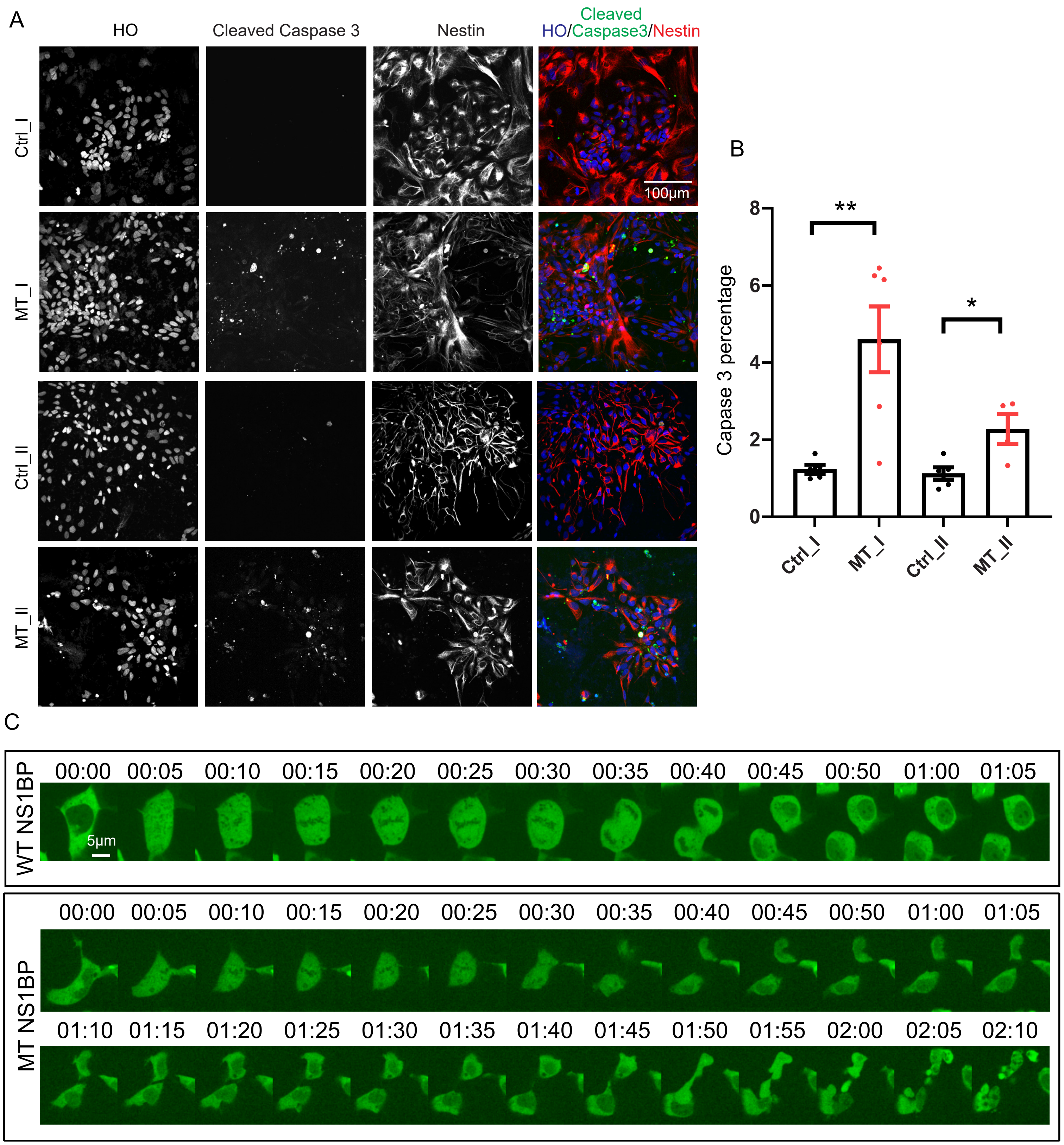


A. Resprentative images of cleaved caspase 3 and nestin staining in the isogenic pairs of NPCs.
B. Quantification in A (Cell numbers >=500 in each group).
C. MT NPCs underwent cell death after mitotic phase while WT NPCs showed normal division.

Figure S6 AP-MS Analysis for the interactors of WT/MT IVNS1ABP

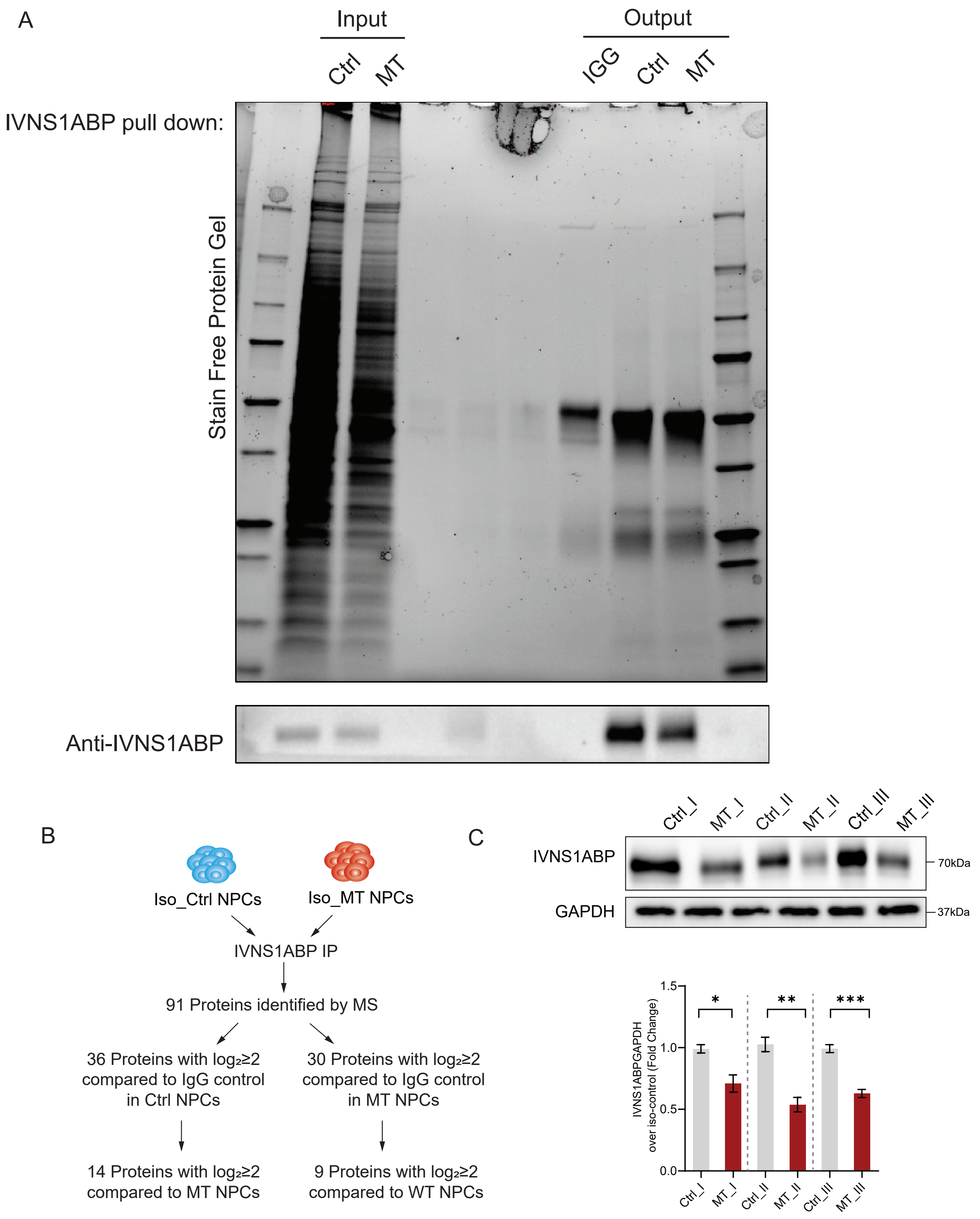


A. Whole gel image showing the successful pull-down of IVNS1ABP.
B. Schematic flow for identifying the interactors of WT/MT IVNS1ABP.
C. Immunoblot and quantification for IVNS1ABP in NPCs derived from isogenic pair I, II & III. n≥3. *, $p \le 0.05$; **, $p \le 0.01$ (One-way ANOVA).

Figure S7 IVNS1ABP mutation leads to the reduced F-Actin binding affinity.

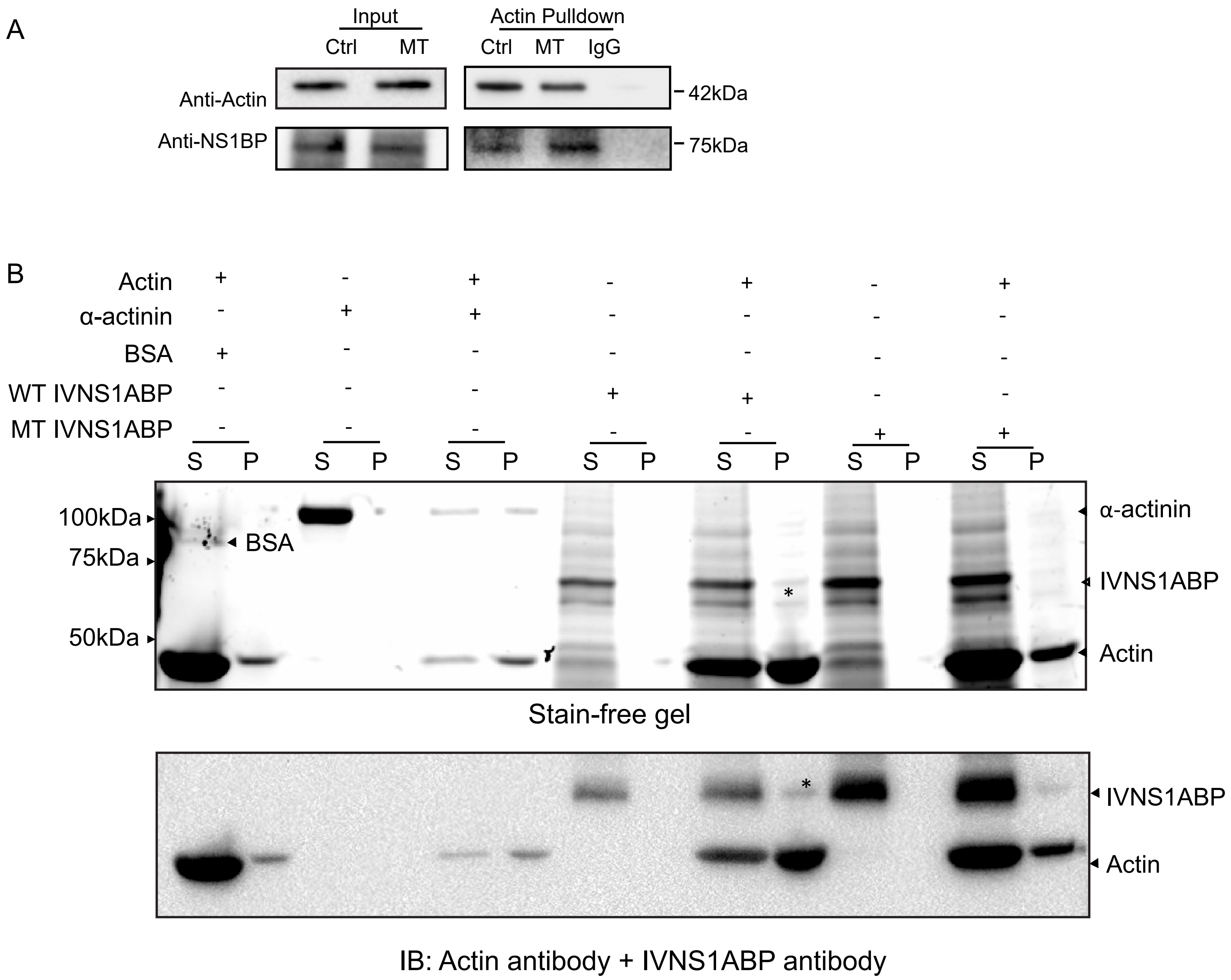


A. Actin pulldown reveals IVNS1ABP expression in both Ctrl and MT group NPCs. IgG serves as negative control.
B. Actin binding assay showed WT IVNS1ABP could bind with G-actin and be co-sedimentated together with F-actin while MT IVNS1ABP showed reduced actin binding affinity. BSA served as negative control and α-actinin served as positive control. S, supernatant (G-actin), P, pellet (F-actin). * represented successful co-sedimentation of WT IVNS1ABP. Up: Stain-free gel image; below: Immunoblotting image of Actin antibody and IVNS1ABP antibody.

Figure S8 Actin is prone to depolymerization by IVNS1ABP mutation

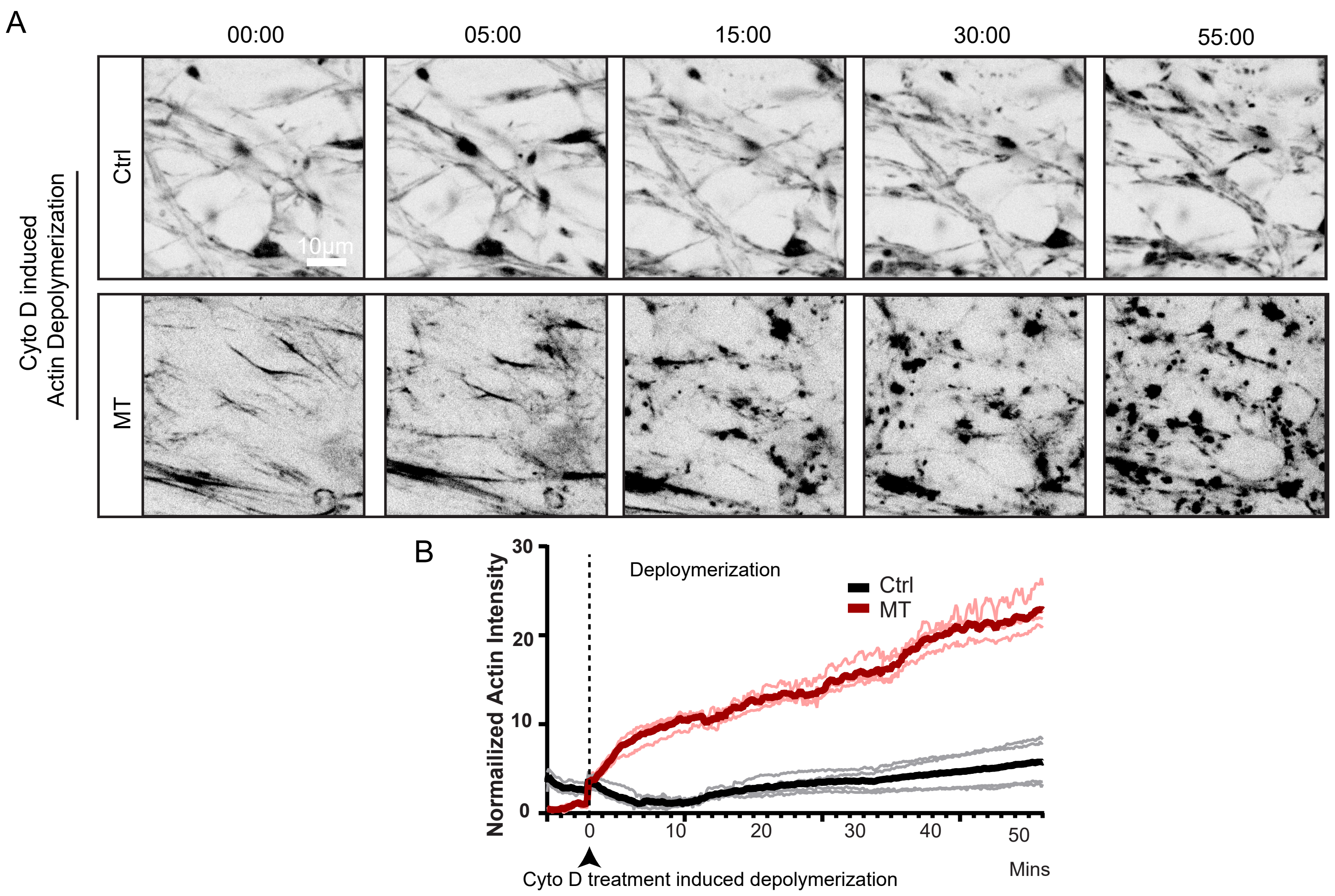


A. Time-lapse of Cytochalasin D (2uM) induced actin depolymerization between Ctrl and MT neural progenitor cells.
B. Quantification of Actin intensity in B (n=3 independent experiments).
